# Rare Maternally Inherited Coding Variants on Chromosome X Carry Predominantly Male Risk in Autism, Tourette Syndrome, and Attention-deficit/Hyperactivity Disorder

**DOI:** 10.1101/2022.09.22.22280248

**Authors:** Sheng Wang, Belinda Wang, Vanessa Drury, Sam Drake, Nawei Sun, Hasan Alkhairo, Juan Arbelaez, Clif Duhn, Tourette International Collaborative Genetics (TIC Genetics), Vanessa H. Bal, Kate Langley, Joanna Martin, Jinchuan Xing, Gary A. Heiman, Jay A. Tischfield, Thomas V. Fernandez, Michael J. Owen, Michael C. O’Donovan, Anita Thapar, Matthew W. State, A. Jeremy Willsey

**Author notes:** Please address correspondence to (A. J. W.).

## Abstract

Autism spectrum disorders (ASD), Tourette syndrome (TS), and attention-deficit/hyperactivity disorder (ADHD) display strong male sex bias, due to a combination of genetic and biological factors, as well as selective ascertainment. While the hemizygous nature of chromosome X (Chr X) in males has long been postulated as a key point of “male vulnerability”, rare genetic variation on this chromosome has not been systematically characterized in large-scale whole exome sequencing studies of “idiopathic” ASD, TS, and ADHD. Here, we take advantage of informative recombinations in simplex ASD families to pinpoint risk-enriched regions on Chr X, within which rare maternally-inherited damaging variants carry substantial risk in males with ASD. We then apply a modified transmission disequilibrium test to 13,052 ASD probands and identify a novel high confidence ASD risk gene at exome-wide significance (*MAGEC3*). Finally, we observe that rare damaging variants within these risk regions carry similar effect sizes in TS and ADHD, further clarifying genetic mechanisms underlying male vulnerability in multiple neurodevelopmental disorders that can be exploited for systematic gene discovery.

## INTRODUCTION

Many neurodevelopmental disorders (NDD), including autism spectrum disorder (ASD), Tourette syndrome (TS), and attention-deficit/hyperactivity disorder (ADHD), have consistent and pronounced male sex biases^1–3^. This male sex bias remains largely unexplained. While ascertainment methods and potential diagnostic bias are likely confounds^1, 4^, studies that endeavored to account for these factors have nonetheless observed residual evidence for male bias^5–7^.

A potential explanation for this remaining male predominance could be the so-called “female protective effect” (FPE), which may be mediated by sex-differential biological factors, such as sex hormones and/or underlying differences in development and physiology^5^. Consistent with this hypothesis, genetic studies of rare de novo and transmitted variants show an increased burden in female probands^8–10^. Similarly, common variants are overrepresented in female probands as well as in unaffected mothers^11–13^.

Along these lines, differences in the canonical composition of the sex chromosomes may also contribute to female “resilience” (or male “susceptibility,” depending on perspective). For example, the presence of a single copy of chromosome X (Chr X) in males likely results in a corresponding susceptibility to deleterious genetic abnormalities, especially within the non-pseudoautosomal region (Chr X non-PAR).

Indeed, genetic disruptions of Chr X have long been studied in psychiatric syndromes and NDDs^14–17^. Over a hundred genes have been associated with X-linked monogenic disorders that predominantly affect males, and are often characterized by severe intellectual disability (ID), structural brain abnormalities, and/or epilepsy^16, 18–20^. Interrogation of rare and severe syndromes with a highly characteristic presentation and substantial comorbidity with ASD, ADHD, epilepsies, ID, and other psychiatric and NDDs has also identified specific X-linked genetic risk factors. These include Chr X aneuploidies such as Turner syndrome and Klinefelter syndrome^15, 21–25^; and disruptions of single genes on Chr X, such as *FMRP*, *MECP2*, *DMD*, and many others^18, 26–32^.

However, systematic, exome- and genome-wide studies of “idiopathic” forms^11^ of ASD, TS, or ADHD have been less successful in identifying risk genes on Chr X^10, 33–35^, especially compared to the hundreds of risk genes that have been identified on the autosomes in these studies^8, 10, 36–46^. *NLGN3* and *NLGN4,* the earliest replicated genes discovered in non-syndromic ASD, identified through mapping cytogenetic abnormalities or performing parametric linkage analysis followed by targeted sequencing, map to Chr X^24, 47–50^. Analyses of structural variation on Chr X have also identified putative risk regions and genes (e.g. Xp22.1 / *PTCHD1-PTCHD1AS*)^51–53^. In these cases, many risk variants are penetrant clinically only in males and have been found to be inherited from unaffected carrier mothers^48, 51^.

Within whole exome sequencing case-control data, Lim *et al*. (2013) previously observed that rare Chr X hemizygous nonsense and canonical splice-site variants were significantly enriched in male ASD probands whereas the corresponding heterozygous variants were not enriched in female probands^54^. Their findings suggested that hemizygous variants within Chr X non-PAR might carry male-specific risks and potentially explained a small proportion of male sex bias in ASD -- though this was not quantified systematically and the female sample size in their study was relatively underpowered to detect such an effect. Additionally, the contribution of Chr X non-PAR missense variants to ASD was not assessed and specific risk genes were not identified.

Since rare likely gene-disrupting (LGD) variants (specifically nonsense, frameshift, and canonical splice-site altering mutations) and missense variants on the autosomes carry well-replicated risks in ASD^8, 10, 42^, it stands to reason that rare missense variants on Chr X may carry risk as well but that the signal may have been obscured by a relative lack of power. While increasingly large cohorts of patients with ASD have been sequenced (e.g.^10, 46, 55^), this question has not yet been resolved, and therefore, current estimates of the contribution of Chr X non-PAR variants to male sex bias in ASD are notably incomplete, especially as missense variants occur much more frequently than LGD variants^10^. Likewise, these questions have not been addressed in other NDDs with pronounced male-bias, such as TS or ADHD, due to limited sample sizes and insufficient power. Finally, combining autosomal LGD and missense variants has been a highly successful strategy for systematic gene discovery in ASD, TS, and other neurodevelopmental and psychiatric disorders^8, 16, 33–35, 42, 56–58^. Consequently, the addition of systematic, reliable analyses of a broad range of rare coding variants mapping to Chr X non-PAR would also be expected to improve the yield of risk gene identification on Chr X.

Here, we analyzed whole exome sequencing (WES) data from male and female ASD probands, leveraging the family-based study design of the Simons Simplex Collection (SSC) to identify rare, maternally-inherited variants on Chr X (**Figure 1**). We focused on maternally-inherited variants for several reasons. First, we hypothesized that the haploid nature of Chr X in males results in a corresponding vulnerability to hemizygous variants that is not present or greatly reduced in heterozygous females. Hence, in simplex families–wherein, by definition, both parents are unaffected–we further hypothesized that some unaffected mothers may be carrying deleterious Chr X variants penetrant predominantly in males, and therefore, that maternally-inherited variants would be enriched in male but not female probands. This is consistent with previous work suggesting transmission of deleterious variants from “carrier” mothers to affected offspring^59^. Second, maternally-inherited variants can be called with high sensitivity and specificity since identifying rare heterozygous variants in diploid mothers is routine with current state-of-the-art strategies^60^. In contrast, *de novo* variants on Chr X are exceedingly rare and technically challenging to call^10, 60, 61^; thus downstream analyses would be fraught with power issues^10^.

**Figure 1.**
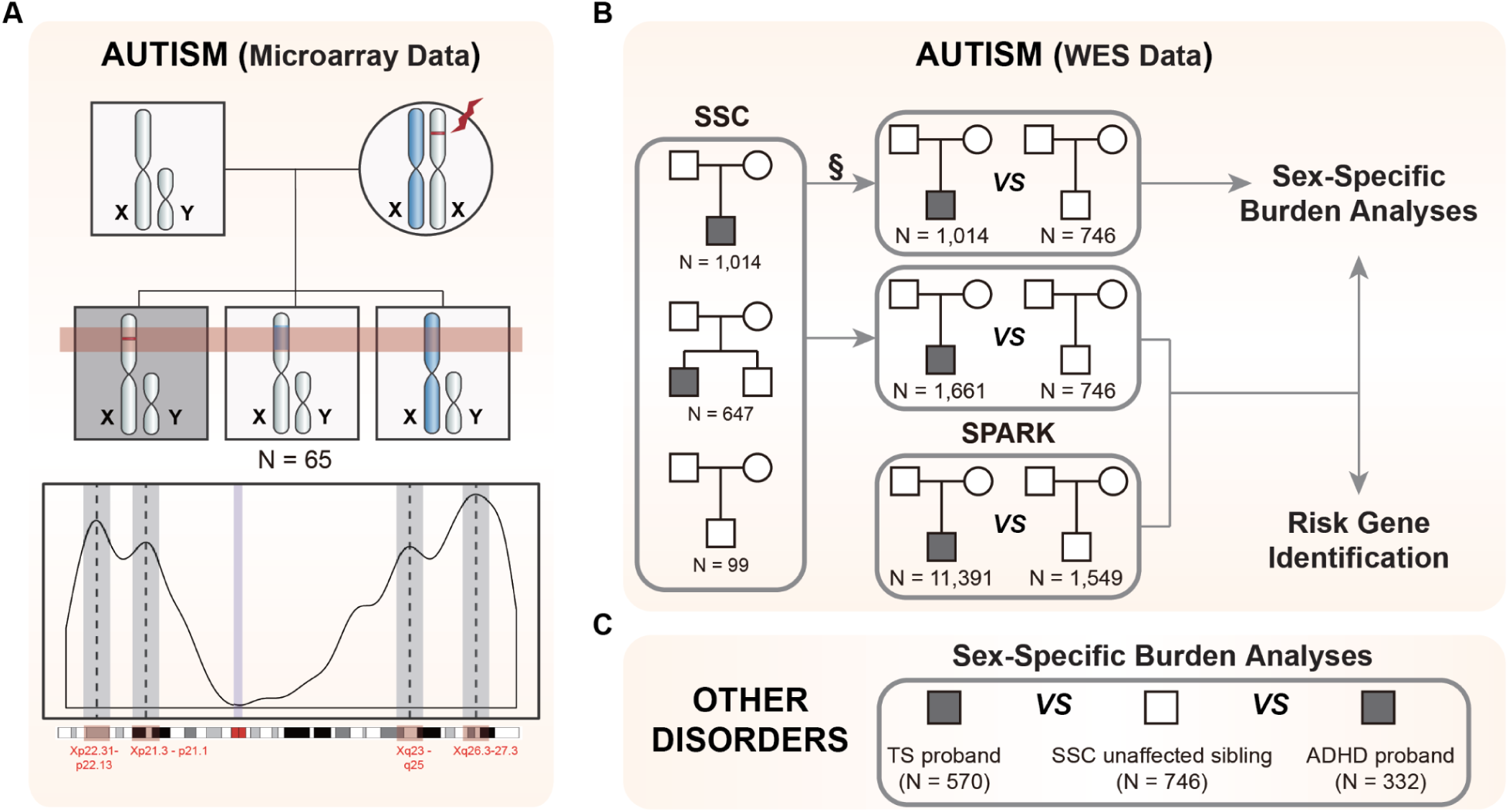
Study schema. (A) We identified risk-enriched regions (RERs) on Chromosome X (Chr X) using microarray data from 65 quintets, consisting of two unaffected parents, one male ASD proband (box in dark) and two unaffected male siblings, where at least one of the unaffected siblings shares the same Chr X origin as the proband (top panel). We identified 4 peaks (RERs) within Chr X non-pseudoautosomal regions (Chr X non-PAR), encompassing a total of 149 genes (bottom panel). (B) We then utilized published whole-exome sequencing data from the SSC and SPARK ASD cohorts for (1) sex-specific burden analyses (SSC for primary analyses, SPARK for validation, combined cohort for final estimation of effect sizes) and (2) risk gene identification (SSC & SPARK combined). For burden analyses, (1) we leveraged SSC siblings as controls as they are well-characterized and do not have reported psychiatric or developmental disorders. §: Chr X data is not independent for SSC families with both male proband and male sibling, and therefore, we trimmed the male probands from such families. We kept the male control siblings because they are the more limited sample set. We similarly trimmed individuals from families with multiple female children, though in this case we removed female unaffected siblings because female probands are more limiting. (2) We combined SSC and SPARK samples to investigate whether rare damaging variants are over-transmitted in male probands. In this analysis, as the untransmitted variants in each individual serve as controls, we include all individuals from SSC cohorts. Female probands are still rare even after combining two cohorts and not with sufficient power for this analysis. For risk gene identification, we integrated all SSC and SPARK male samples with maternal data. (C) We extended our analyses to TS and ADHD in order to determine whether RER scarry risk in other male-biased psychiatric disorders.

Comparing 1,014 male ASD probands to 746 male siblings from the SSC, we confirm the previously demonstrated overrepresentation of rare maternally-inherited Chr X non-PAR LGD variants in male cases but do not observe evidence for the contribution of probably damaging “missense 3” (Mis3) variants alone (PolyPhen2 [HDIV] score ≥ 0.957^62, 63^) or of rare damaging variants (LGD + Mis3) as a group. To better stratify risk-carrying variants, we leveraged microarray genotyping data from SSC families with multiple male children to identify specific regions within Chr X non-PAR that consistently segregated with risk–an approach conceptually similar to the newly developed stratified polygenic transmission disequilibrium test^64, 65^ (**Figure 1**). Strikingly, within these regions both Mis3 variants alone as well as LGD + Mis3 variants as a group (i.e., damaging variants) showed highly significant enrichment. We then replicated this observation by demonstrating transmission disequilibrium of both LGD and Mis3 variants in males from the SPARK ASD cohort^66^. Next, we combined 1,661 SSC male probands with an additional 11,391 SPARK male probands and utilized a novel modified transmission disequilibrium test to pinpoint one exome-wide significant ASD risk gene (*MAGEC3*) (**Figure 1**).

Finally, we reproduced this analytic approach in TS (N = 570) and ADHD (N = 332) WES datasets (**Figure 1**) and observed robust evidence for Chr X risk in males for these two strongly sex-biased psychiatric disorders, but not for epileptic encephalopathies (EE, N = 223), which affect males and females at a similar frequency^67–69^--suggesting that susceptibility to hemizygous damaging variants is a common mechanism in NDDs with male sex bias and that their large-scale identification offers a powerful addition to the armamentarium for systematic gene discovery in these disorders.

## RESULTS

### Rare transmitted damaging variants are not enriched Chromosome-X-wide in ASD

We first analyzed WES data from 2,058 simplex ASD families (7,771 samples), including 1,597 quartets and 461 trios from the SSC, representing 1,975 probands and 1,680 unaffected siblings (**Table S1**)^8, 35, 42^. For all burden analyses, we used unaffected siblings as controls. However, as proband-sibling pairs from the same family are not independent (i.e. siblings could share the same Chr X), we selected either one proband or one sibling from each quartet family, prioritizing male controls and female probands as these were the most limiting sample sets. This resulted in 1,328 ASD probands (males: 1,014, females: 314) and 1,557 unaffected siblings (males: 746, females: 811) (**Figure 1**).

We focused on rare (minor allele frequency (MAF) ≤ 0.1% in ExAC v0.3 and ≤ 0.1% within the SSC dataset), maternally-inherited, hemizygous coding variants on the non-PAR of Chr X (808 genes) in male probands and SSC male control siblings. We normalized the mutation rate by the rate of rare synonymous variants in order to control for differences in sequencing platforms and ancestry (**Table S1**, **Figure S1**). We then compared the rate of hemizygous LGD variants in male ASD probands versus male control siblings and observed enrichment consistent with previous reports^46, 54^ (OR 1.86, p = 0.028, one-sided Fisher’s exact test; **Table S2**). We did not observe enrichment of Mis3 variants alone (OR 0.94, p = 0.78) or of all damaging variants (LGD + Mis3) when analyzed together (OR 0.97, p = 0.65)--again, consistent with the previous work^46, 54^. We also did not observe significant enrichment in female ASD probands versus female control siblings for any of these variant classes (**Table S2**).

### Rare transmitted damaging variants are enriched in defined risk-enriched regions in males with ASD

Given replicated evidence for the contribution of transmitted LGD mutations on Chr X, but not for missense or damaging mutations as a group, we reasoned that strategies similar to those employed on the autosomes to stratify risk alleles, for example restricting to constrained genes, might improve signal detection and enhance gene discovery. However, constraint metrics are estimated based on selection pressures for a diploid genome, which may be different for Chr X. Consequently we investigated whether restricting the search space to Chr X regions over-transmitted from mothers to affected sons might lead to improved detection of risk alleles. This is conceptually similar to a recently developed approach leveraging common variant polygenic risk scores from the autosome to identify blocks of excess over-transmission of ASD polygenic risk (so-called stratified polygenic transmission disequilibrium test or S-pTDT)^64, 65^.

We turned to microarray genotyping data from SSC families consisting of a male proband and at least two unaffected male siblings (n = 65) to identify regions on Chr X non-PAR that segregated uniquely to the male proband within a given family, with the expectation that these regions would be enriched for genes carrying risk (**Figure 1A, Figure S2A**). We term these “risk-enriched regions” (RERs) and denote the remainder of Chr X non-PAR as non-enriched regions (NERs). There are 149 genes within the RERs and 659 genes in NERs.

Within the RERs, damaging variants as a group are now significantly enriched in male probands (OR 1.60, p = 0.0084, one-sided Fisher’s exact test). This signal is specific to RERs and absent for more common variants (MAF > 0.1%) (**Figure 2A**). Moreover, our model posits that females are protected from rare heterozygous mutations, and consistent with this hypothesis, rare maternally transmitted heterozygous damaging variants in female probands are not enriched within the RERs (OR 1.11, p = 0.39) (**Figure 2A**).

**Figure 2.**
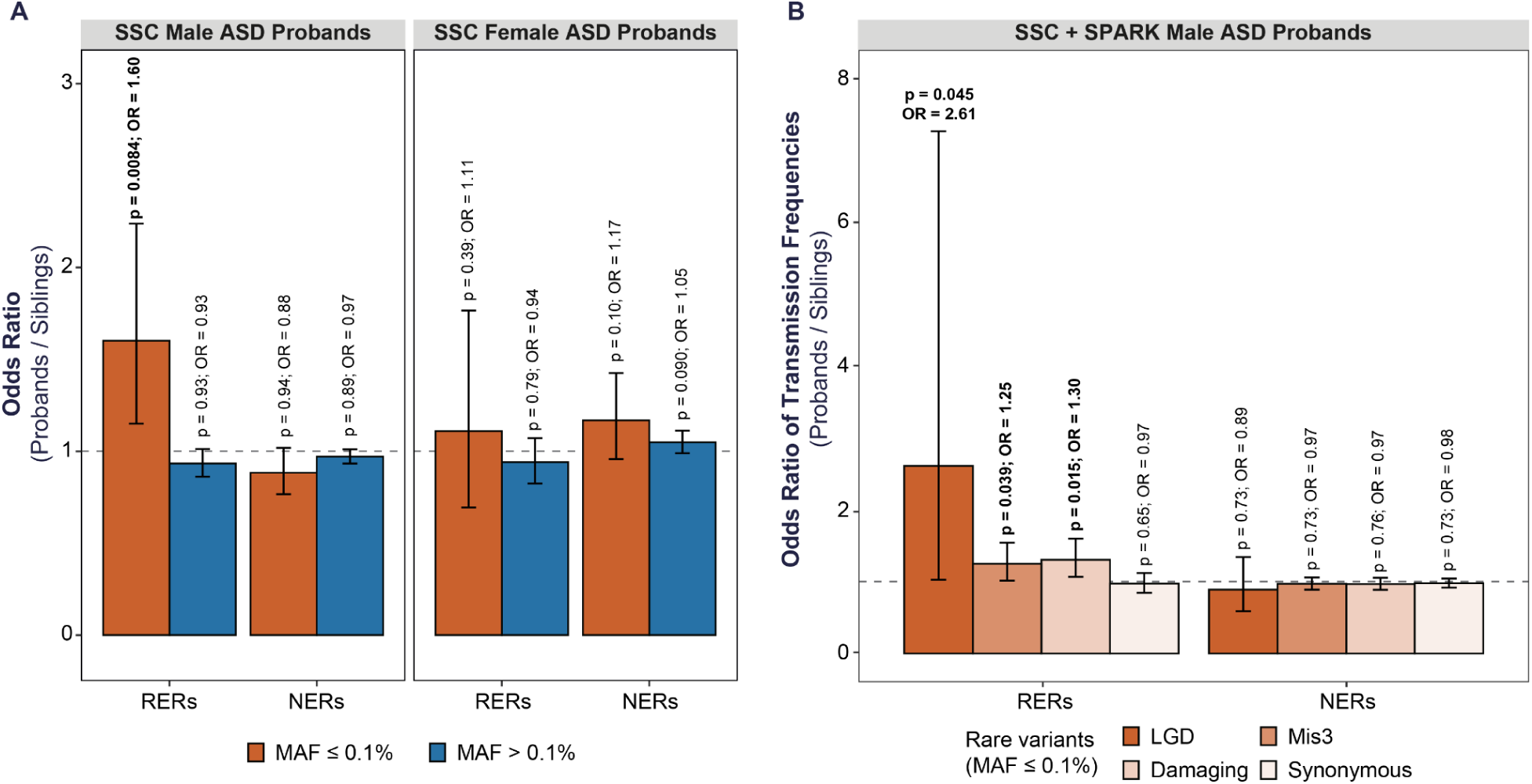
Rare transmitted damaging variants are enriched in RERs. We defined risk-enriched regions (RERs) based on patterns of segregation in a microarray dataset from the SSC (see Figure 1) and considered any regions outside of the four risk regions to be non-enriched regions (NERs). We then compared the rate of (maternally) transmitted damaging variants in probands versus SSC siblings, utilizing the rate of synonymous variants to control for potential differences in sequencing metrics and ancestries (see related **Figures S1-2**). (A) Rare (MAF ≤ 0.1%) transmitted damaging variants are enriched in RERs in male but not female probands. Rare damaging variants in NERs are not enriched in males or females, nor are more common variants (MAF > 0.1%) enriched in RERs or NERs. (B) We further confirmed the enrichment of rare damaging variants in male probands from SSC and SPARK families by comparing the transmission probabilities of rare variants. Separately, LGD and Mis3 variants are also overtransmitted. The gray horizontal line indicates OR = 1 and 95% confidence intervals are shown in black error bars. Damaging variants consist of likely gene disrupting (LGD) and missense 3 (Mis 3; PolyPhen2 [HDIV] score ≥ 0.957) variants.

Additionally, we reasoned that risk may be increased within genes highly expressed in the brain, as has been shown for autosomal genes in ASD^38^. We leveraged BrainSpan^70, 71^ pre- and post-natal male gene expression data from 16 regions of the human brain to rank genes by overall expression. We observed the same phenomenon within RERs: rare transmitted damaging variants impacting male probands are significantly enriched only in the subset of RER genes that rank among the top 50% of brain expressed genes (OR 2.10, p = 0.015, expression ranking is relative to the entire exome), though some signal is observed in the subset of RER genes ranking in the bottom 50% (OR 1.50, p = 0.085). Again, this is a male-specific effect (females: OR 1.07, p = 0.53, **Figure S2B**).

De novo autosomal variants are associated with decrements in non-verbal IQ (NVIQ)^8, 10, 42^. We therefore assessed whether rare damaging variants within RERs carry a similar pattern. We first compared NVIQ in male probands with damaging RER variants to those without and observed no clear difference (mean NVIQ 87.56 versus 86.27, p = 0.31). We next split the SSC cohort by NVIQ into two groups, one below the cohort average (NVIQ < 85; n = 389 male probands) and the other average or above for the cohort (NVIQ ≥ 85; n = 593 male probands). We did not observe an increase in rare damaging variants in the below average NVIQ group. In fact, the enrichment of rare damaging variants may be higher in the above average NVIQ group (OR 1.40 p = 0.10; **Figure S2C**). Taken together, these results suggest that RER risk variants do not appear to be associated with decrements in NVIQ, unlike autosomal de novo variants^42, 72–74^.

Next, we conducted several analyses to determine whether our results could be explained by a range of confounds including our normalization method or population stratification. First, we compared the unnormalized mutation rates between probands and siblings and observed the same male-specific signal for rare variants (Rate ratio 1.36, p = 0.034, one-sided Poisson test; **Figure S3A**). Subsequently, we narrowed to the subset of SSC probands and siblings with European ancestry and repeated our burden analyses in males and females. We observed a remarkably similar and significant effect size for rare damaging variants in European male probands (OR 1.63, p = 0.021; **Figure S3B**) and the same absence of signal in European females (OR 0.75, p = 0.84). Last, we validated our results in an independent dataset and with a different burden test. Specifically, we compared the transmission probability of rare variants within SPARK families (11,391 SPARK male probands, 1,549 male sibling controls)^66^, and observed that only rare damaging variants in RERs are over-transmitted to male probands (OR 1.31, p = 0.014) and that rare synonymous variants in RERs are not over-transmitted to male probands (**Figure S3C**). Taken together, these results suggest that the enrichment of rare damaging variants within the identified RERs are male-specific, not driven by normalization methods, variant calling approaches, systematic batch effects, or ancestry, and apply to ASD cohorts with different ascertainment criteria.

Finally, we combined all the male samples from the SSC and SPARK cohorts (13,052 male probands, 2,295 male sibling controls) and assessed LGD and Mis3 variants separately. Within this large cohort, we identified significant over-transmission of both LGD (OR 2.61, p = 0.045) and Mis3 variants (OR = 1.25, p = 0.039) as well as damaging variants as a group (OR 1.30, p = 0.015). Again, we did not observe signal for any of these variant types in NERs (**Figure 2B**).

### RERs are correlated with recombination hotspots

Our strategy to identify RERs depends on recombination, and therefore, regions with higher recombination rates may be more likely to be identified as RERs. Indeed, the “risk” curve based on segregation in SSC families is highly similar to a curve generated solely from the recombination rates reported for Chr X as a part of the HapMap project^75^ (Wilcoxon rank sum test p = 0.98; **Figure 3A**). To examine this overlap at the gene level, we compared the 149 genes within RERs to the 149 genes with the highest surrounding recombination rates in HapMap (“recombination” gene set). Though the 149 RER genes are enriched for genes with high recombination rate (fold-enrichment 1.81, p = 1.0E-5, permutation test), they occupy a much broader distribution of recombination rates (**Figure 3B-C**). Indeed, only 50 genes of the 149 RER genes (33.6%) are present in the “recombination” gene set, suggesting that these two gene sets may have different patterns of risk. Therefore, we compared the enrichment of maternally-inherited rare damaging variants within the 99 genes unique to the RERs, the 50 overlapping genes, and the 99 genes unique to the recombination gene set. We observed the strongest enrichment within the 50 overlapping genes (OR 1.75, p = 0.028), followed by the 99 unique RER genes (OR 1.48, p = 0.085), and then the 99 unique recombination genes (OR 1.07, p = 0.38). In fact, rare damaging variants are significantly more likely to occur within the 99 RER-only genes than within the 99 recombination-only genes (RR = 1.48, p = 0.036, one-sided Poisson test). Together, this suggests that the majority of risk is present within the RER gene set and that our analysis identified these RERs based on more than just recombination rate alone.

**Figure 3.**
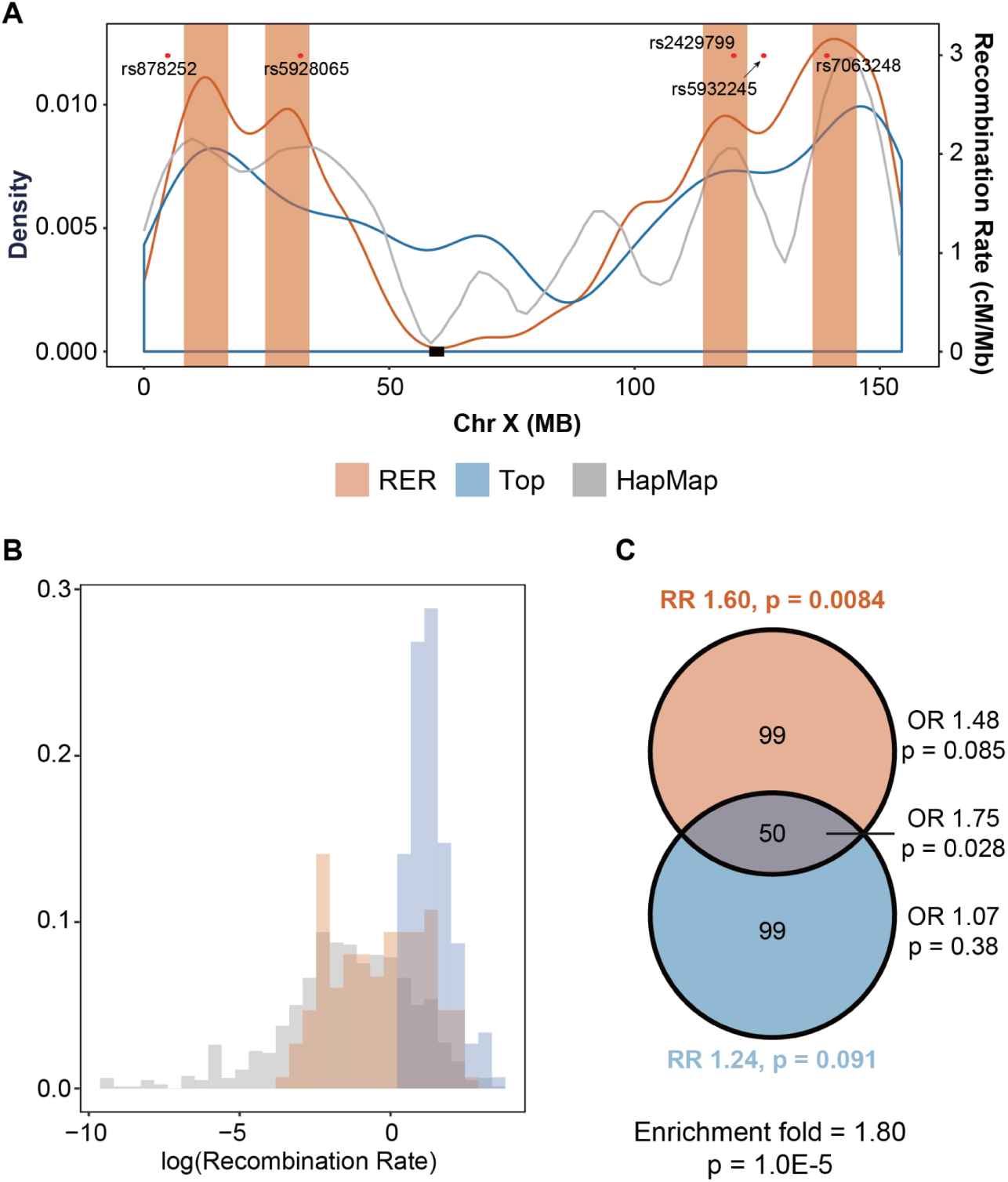
RERs are correlated with local recombination rates. (A) Density curve for risk-enriched regions (RERs) (red), and genes with the highest recombination rate (blue, generated from HapMap). The overall recombination rates from HapMap project^75^ are indicated with a gray smooth line. The red dots correspond to the top five SNPs identified in a Chr-X-wide association study for loci contributing to the female protective effect^85^. (B) RER genes (red) tend to have high rates of recombination compared to all Chr X non-PAR genes (gray). However, the RER genes only partially overlap the distribution of the top 149 genes based on recombination rate (blue). (C) Venn diagram depicting the overlap between the 149 genes contained within RERs and the 149 genes with the highest recombination rate. These two gene sets significantly overlap (permutation test with 100,000 iterations), but most of the risk for rare transmitted damaging variants resides within RER genes (Fisher’s exact tests).

### Meta-analysis with SPARK data identifies *MAGEC3* as a high confidence ASD risk gene

We next sought to identify specific risk genes on Chr X based on an overrepresentation of damaging variants in male probands, as has been done highly successfully for autosomal genes in ASD and other neurodevelopmental and psychiatric disorders^11, 33, 76^. We combined samples from the SSC and SPARK datasets, yielding a total of 13,052 male probands and 2,295 controls (see **Figure 1**, **Figure S4**, **Methods**). Compared to de novo variation, gene discovery using transmitted variation is much more vulnerable to differences in ancestry between cases versus controls^57^. Therefore, we designed a modified transmission disequilibrium test (TDT), as these types of tests are more robust to population stratification because the non-transmitted variants are in effect an ancestry-matched control population^77, 78^. However, one of the challenges in applying rare variant TDTs is the systematic undercalling of rare variants, which shifts the null hypothesis for transmission (detection) below 50%, thereby reducing power for detection of significant overtransmission^78^. Here, we observe even more substantial undercalling of rare maternally-inherited variants on Chr X as compared to rare autosomal inherited variants (**Table S4**). To account for this systematic bias, we therefore estimated null transmission probabilities for each gene based on data from control samples. Since there are a relatively small number of rare variants per gene in control samples, we therefore estimated the local transmission probability for each gene using 3 MB bins (**Methods**). We then conducted the TDTs with these modified transmission probabilities.

Within the 149 genes in the refined RERs and using a threshold of Chr-X-wide significance (p < 6.2E-5 after Chr-X-wide Bonferroni correction for 808 genes), we identified a single gene–*MAGEC3*–with significant over-transmission of rare damaging variants to ASD probands versus male sibling controls (p = 2.10E-07; **Figure 4**, **Table S1**). This gene also passes exome-wide significance (p < 2.5E-06). As a control, we tested NER genes for association, and consistent with the hypothesis that these regions are depleted of risk, we did not identify any genes associated at Chr-X-wide significance within the refined NERs (**Figure 4**).

**Figure 4.**
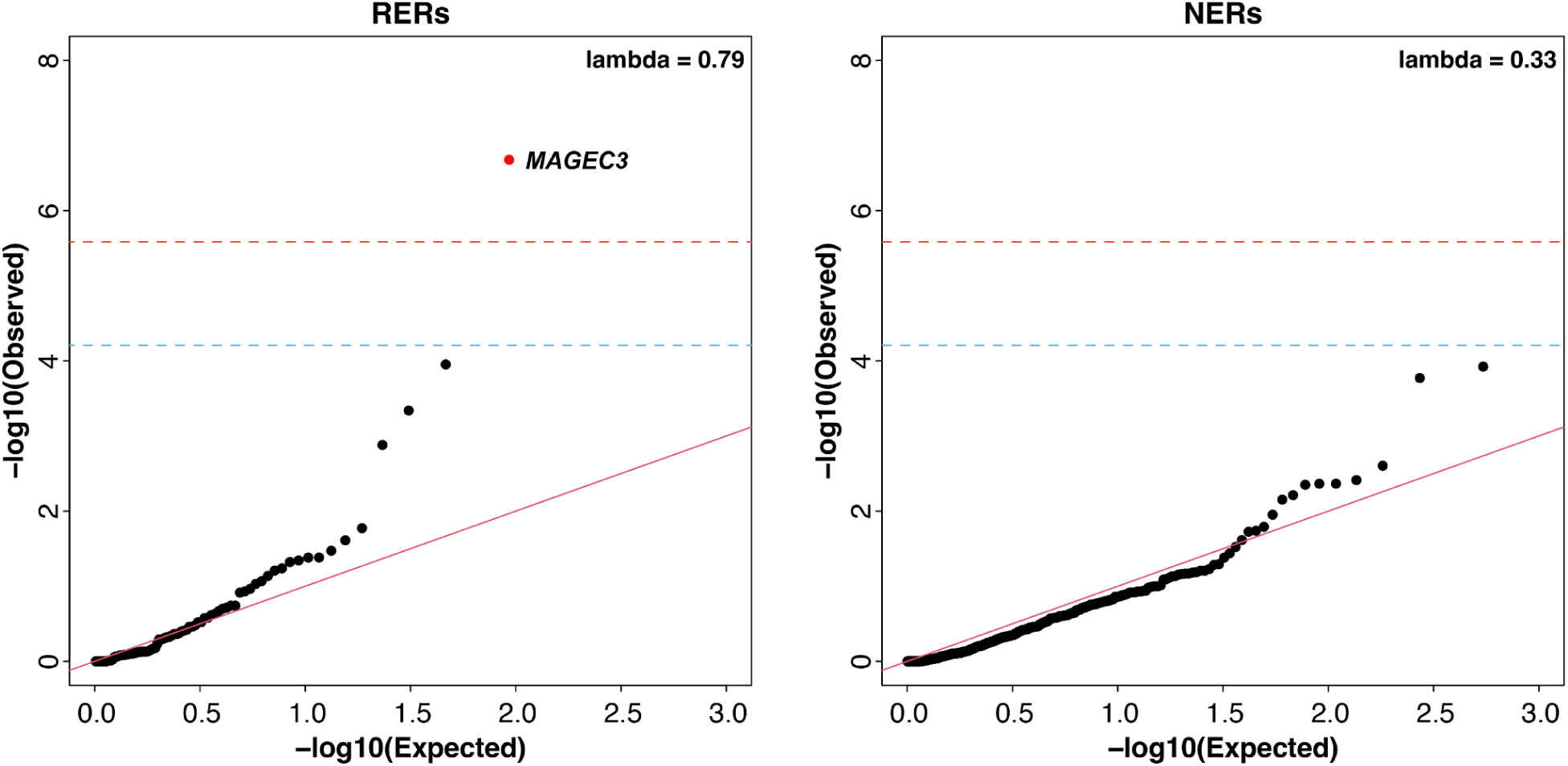
QQ plots for gene discovery. We conducted gene discovery within the refined RERs using a modified transmission disequilibrium test for rare damaging variants. As a control, we also conducted gene discovery in the refined NERs. In each case, we compared the distribution of p values to a uniform distribution (red diagonal line). Chr X non-PAR and exome wide significances are indicated with blue and red horizontal dashed lines, respectively.

### Rare damaging variants in Chr X RERs are enriched in other male-biased neurodevelopmental disorders

Prior studies have demonstrated that ASD shares genetic risk with other disorders, such as ADHD, TS, and EE^33, 79–82^. However, only TS and ADHD are strongly male sex biased^2, 3, 67–69^. We therefore assessed whether rare damaging variants within RERs also contribute risk to these disorders, with the hypothesis that TS and ADHD will carry risk in these regions but that EE will not. We analyzed 570 TS male probands, 332 ADHD male probands, and 223 EE male probands, using the same 746 SSC male siblings as controls. For TS, we utilized 546 previously sequenced male probands^34, 35^ along with 24 newly sequenced male probands; for ADHD, we used 332 newly sequenced male probands; and for EE, we leveraged 223 previously sequenced male probands^82^ (see also **Methods**). We did not generate exome sequencing data for the parents of the 332 ADHD probands. Therefore, to fairly compare the three disorders, for all cohorts we performed analyses using rare hemizygous variants called based on proband data alone (i.e., we did not verify transmission status and instead conducted case-control burden analyses).

Before conducting these analyses, we conducted several analyses to assess the viability of calling rare hemizygous variants in male samples without utilizing maternal data. First, using data from SSC male children only, we estimated our ability to re-identify previously called maternally-inherited damaging variants. We observed a recall rate of 97.24% and a precision of 91.13%. Second, since parental whole exome data is available for the TS probands, we analogously assessed recall rate and precision using mother-son data from the TS cohort (n = 570 pairs). We observed a similarly high recall rate and precision (96.90% and 91.15% respectively). Third, after calling rare hemizygous variants in the SSC male samples, we observed that rare damaging variants are significantly enriched in SSC male cases versus controls with a remarkably consistent effect size (OR 1.45, p = 0.027 for case-control versus OR 1.60, p = 0.0084 for maternally transmitted; see **Figure 5A** versus **Figure 2A**). Finally, we attempted Sanger sequencing based confirmation of all rare coding variants identified in the ADHD samples and observed a high confirmation rate (∼89-98%, see **Methods**). Together, these analyses support the viability of case-control analyses thereby enabling a more direct comparison of the effect sizes in ASD, TS, ADHD, and EE.

**Figure 5.**
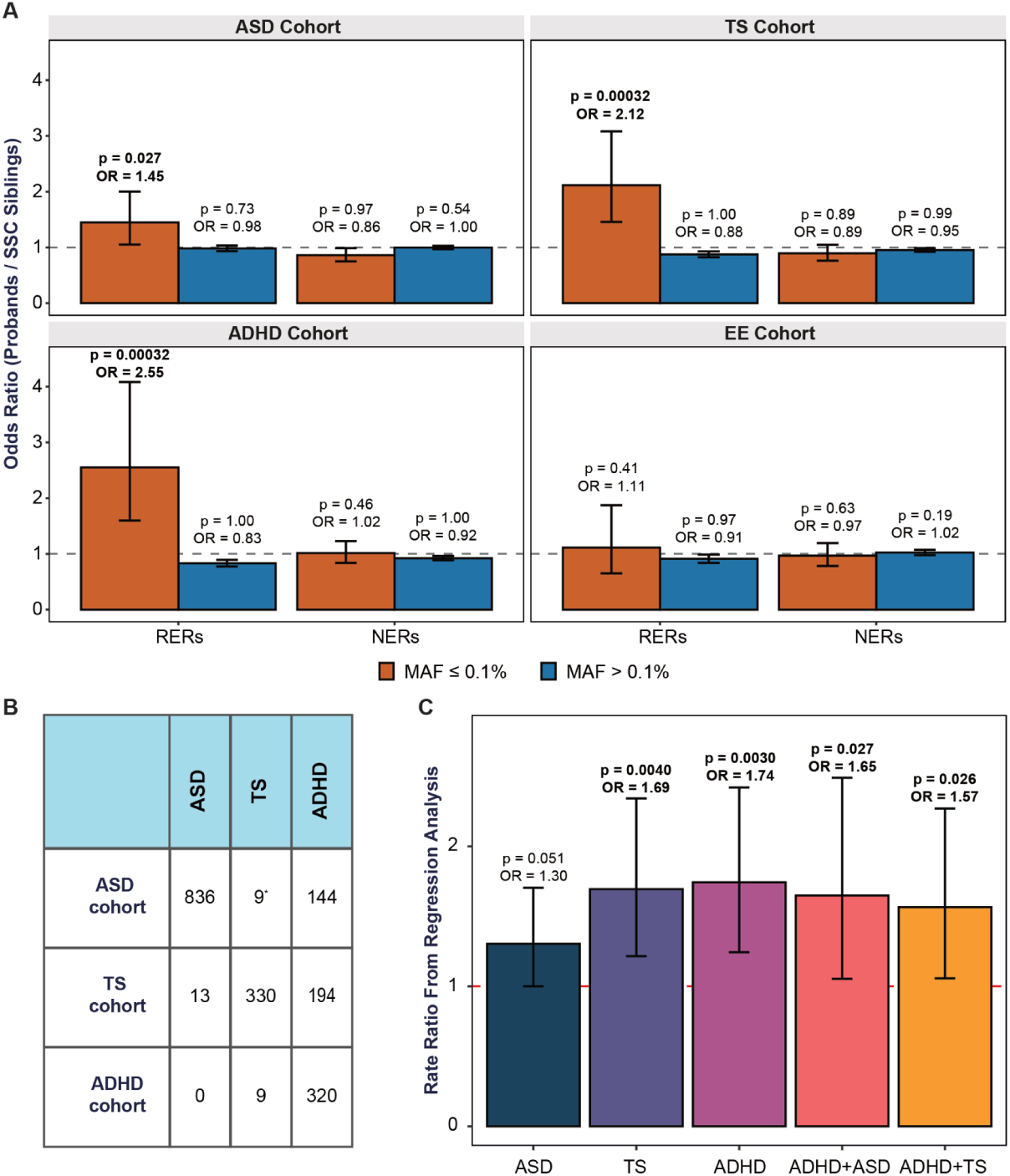
Rare damaging variants in risk-enriched regions (RERs) are also enriched in males with Tourette Syndrome or ADHD. We identified rare damaging variants based on proband data alone for ASD, Tourette syndrome (TS), attention-deficit/hyperactivity disorder (ADHD), and epileptic encephalopathies (EE). In all burden analyses, we used male SSC siblings as controls. (A) Rare (MAF ≤ 0.1%) damaging variants are enriched within RERs in re-analyzed data from SSC male probands, and in the ADHD and TS cohorts. However, they are not enriched in EE, which has minimal sex bias, nor are they enriched in NERs in any disorder. Likewise, more common variants (MAF > 0.1%) are not enriched in any disorder or set of regions. (B) Rare damaging variants appear to be more strongly enriched in male probands in the TS and ADHD cohorts, but this could be due to comorbid TS, ADHD, and/or ASD diagnoses in male probands within each cohort. Only individuals affected by one or two disorders are listed. *In the ASD cohort probands were reported with/without Tourette/Tic disorder whereas in the TS and ADHD cohorts probands were reported with/without TS. (C) We therefore conducted a Poisson regression analysis with ASD, TS, and ADHD status as covariates, the results of which further suggest that male probands with TS and/or ADHD are the most likely to have damaging rare variants in RERs. We excluded patient groups with fewer than 100 samples (e.g. ASD + TS). **See also Table 1, Figure S4**.

**Table 1.** Rare damaging variants in RERs contribute to 2-3% of ASD, TS, and ADHD diagnoses in males. We estimated the contribution of rare damaging variants using variants detected through case-control (ASD, TS, ADHD cohorts) data. According to the ADHD comorbidity status in the ASD cohort, we further conducted the analysis for probands with ASD and ADHD (ADHD+), and probands with ASD but no ADHD (ADHD-). We estimated that more than 2% of male cases can be explained by rare damaging variants in RERs and that approximately 20% of damaging variants carry risk.

| Dataset | Samples | Cases with $\geq 1$ variant (%) | Controls with $\geq 1$ variant (%) | Cases with contribution from $\geq 1$ variant (% with 95% CI) | Variants carrying risk (% with 95% CI) |
| --- | --- | --- | --- | --- | --- |
| Case-control | ASD Cohort | 10.85 | 8.49 | 2.36<br>(0 - 5.17) | 19.68<br>(0 - 46.47) |
|  | ASD Cohort (ADHD+) | 14.00 |  | 5.51<br>(0 - 11.68) | 35.42<br>(0 - 78.51) |
|  | ASD Cohort (ADHD-) | 10.47 |  | 1.98<br>(0 - 4.87) | 17.43<br>(0 - 46.15) |
|  | TS Cohort | 11.23 |  | 2.74<br>(0 - 6.09) | 27.54<br>(0 - 57.68) |
|  | ADHD Cohort | 11.25 |  | 2.75<br>(0 - 6.90) | 23.73<br>(0 - 60.16) |

We next analyzed the TS and ADHD samples and, strikingly, observed that both cohorts are strongly enriched for rare hemizygous damaging variants, with effect sizes larger than we observed in ASD (TS: OR 2.12, p = 0.00032; ADHD: OR 2.55, p = 0.00032, ASD: OR 1.45, p = 0.027; one-sided Fisher’s exact test; **Figure 5A**). As a negative control, we analyzed 223 male probands with EE, again based on the hypothesis that EE cases will not show strong enrichment for rare damaging variants within Chr X RERs due to the relative lack of sex bias in this disorder. Indeed, we do not observe a statistically significant excess in EE cases versus SSC controls (OR 1.11, p = 0.41). All together, this suggests that rare damaging variants within RERs predominantly carry risk for males in male-biased NDDs only.

ASD, TS, and ADHD are commonly comorbid^33, 79–81^. Therefore, to better understand the relative effect sizes for rare hemizygous variants within Chr X RERs, we conducted a Poisson regression analysis with phenotype(s) as a covariate (**Figure 5B-C**). We grouped the samples from each cohort based on comorbidity status and excluded groups with fewer than 100 samples due to a lack of power. Again, rare damaging variants on Chr X appeared to carry more risk in TS (OR 1.69, p = 0.004) and ADHD (OR 1.74, p = 0.003) as compared to ASD (OR 1.30, p = 0.05). We confirmed this result by comparing rare transmitted damaging variants in RERs in SSC probands with elevated ADHD symptoms versus those without (**Methods**). While both groups were significantly enriched for damaging variants on Chr X (ASD only: OR 1.46, p = 0.036; ASD with ADHD: OR 2.64, p = 0.0023), the rate in ASD with elevated ADHD symptoms was significantly greater (OR 1.82, p = 0.041; **Figure S2D**).

Finally, we estimated the proportion of rare damaging variants that carry risk for each disorder and the proportions range from 19.68% (0 - 46.47%) in ASD to 23.73% (0 - 60.16%) ADHD, and 27.54% (0 - 57.68%) in TS (**Table 1**). We also estimated the percentage of cases in which these variants contribute to risk, yielding highly similar estimates of 2-3% (ASD: 2.36% (0 - 5.16%), TS: 2.74% (0 - 6.03%), ADHD: 2.75% (0 - 6.68%). Consistent with our previous observations, in ASD probands from the SSC, the contribution of rare damaging variants in RERs varied depending on the presence of absence of elevated ADHD symptoms: these variants contribute risk in 5.51% of male probands with “comorbid” ADHD versus 1.98% of those without “comorbid” ADHD (**Table 1**, **Figure S2D**). Altogether, these results suggest that rare hemizygous damaging variants within Chr X RERs carry broad risk for male-biased neurodevelopmental disorders and that gene discovery will be viable in larger cohorts of TS and ADHD samples.

## DISCUSSION

In this study, we identified, for the first time, a clear pattern of risk for rare hemizygous damaging variants within specific RERs on Chr X. This signal appears to be male-specific and spans patients with ASD, TS, and/or ADHD--but not EE, an NDD with limited male sex bias. Previous case-control work has shown that LGD mutations on Chr X non-PAR contribute ASD risk to males^54^. We confirm that result here, observing a similar effect size Chr X-wide despite focusing on maternally-inherited variants (**Table S2**). However, we were unable to identify any enrichment for rare Mis3 variants, or damaging variants as a group, until narrowing our variant calls to the 149 genes within the RERs we delineated from analyzing patterns of segregation in simplex SSC families with multiple male children. Enrichment of damaging variants within the RERs persisted with and without normalization, does not appear to be due to population stratification, and replicated in an independent cohort (**Figures S1**, **S3**). Moreover, we observed that the LGD variants identified in males by Lim *et al*. appear to be enriched in the RERs as compared to the rest of Chr X non-PAR (rate ratio 2.15 versus 1.43; OR 1.49, p = 0.37). Similarly, a recent omnibus study of ASD by Zhou *et al*.^46^ identified a significant enrichment of maternally-inherited rare LGD variants in male probands and we observed that in this much larger cohort maternally-inherited rare LGD variants are significantly enriched in RERs compared to NERs (OR 1.25, p = 0.05). However, similar to the Lim *et al*. paper, enrichment of maternally-transmitted missense variants was not reported. Regardless, these observations suggest that our results are not confounded by systematic biases in variant calling or ancestry and that the RERs carry risk in multiple cohorts.

Identification of the RERs relied on ‘informative’ recombination events, therefore it is unsurprising that the RERs positively correlate with local recombination rates (**Figure 3**). While a high recombination rate alone does not identify regions of increased risk as accurately as our model (OR 1.24, p = 0.091 versus OR 1.60, p = 0.0084), we cannot exclude the possibility that there is additional risk carried in regions of high recombination, especially because previous work in ASD has suggested that hotspots of de novo sequence and copy number variation are significantly related to recombination rate^72, 83, 84^.

Interestingly, these four regions appear to co-locate with the top five SNPs from a targeted genome-wide association study (GWAS) for a common genetic locus on Chr X that may mediate the FPE in ASD^85^ (**Figure 3**). While the significance of this is unclear, it suggests that common variants within the RERs may also impact risk/resilience (or tag rare variants/haplotypes that impact risk). As polygenic risk scores for Chr X SNPs are generated, it will be important to address this question more directly. Finally, even though the RERs were identified from an ASD cohort, damaging variants within these RERs also contribute risk in TS and ADHD. This might suggest that these conditions share genetic risk on Chr X, consistent with what has been observed for autosomal rare and common variation, the elevated sex ratio in probands with comorbid ASD and ADHD^33, 35^, and the increased rate of Chr X risk variants in SSC patients with elevated ADHD symptoms (**Figure S2D**).

Strikingly, the effect sizes estimated here parallel or exceed those observed for de novo damaging variants in ASD, TS, and ADHD^8, 33, 35^. However, the percentage of cases in which these variants contribute to risk is substantially lower (2-3% versus > 9%)^8, 33, 35^. This may be due, in part, to the relatively small size of the Chr X RERs compared to the 22 autosomal chromosomes as well as the haploid nature of Chr X in males. In that light, this is a remarkably high contribution from such a small proportion of the exome.

Large-scale sequencing studies have been highly successful in idiopathic ASD–identifying hundreds of autosomal risk genes^8, 10, 36–46^. However, gene discovery on Chr X has lagged behind due to several reasons, including technical difficulties in calling rare variants and the rarity of de novo coding variants, both of which reduce power for gene discovery via TDTs and/or recurrent de novo variation, respectively. Here, leveraging RERs, a combined dataset of 13,052 male probands and 2,295 controls from the SSC and SPARK, and a modified TDT test accounting for undercalling of maternally-inherited Chr X rare variants, we identified a single gene, *MAGEC3*, with exome-wide significant over-transmission of rare damaging variants. *MAGEC3* belongs to the melanoma antigen (MAGE) family. Proteins from this family, which interact with E3 RING ubiquitin ligases to regulate ubiquitination, have been implicated in a wide range of disorders, including NDD^86^. This gene is also known to escape Chr X inactivation in females^87, 88^. Despite the fact that Chr X is enriched for ID genes^16, 18^, to the best of our knowledge, this high confidence risk gene has not been associated with ID. It also does not overlap with the top 23 genes identified in the recent DDD study of Chr X^16^.

Overall, these results raise promising hypotheses about the underlying biology of ASD, TS, and ADHD and argue for additional investigation of genetic risk on Chr X in these and other NDDs with a male sex bias, with the expectation that additional risk genes will be identified as sample sizes increase in ASD, TS, and ADHD. It will also be important to characterize the intersection of this male-specific risk factor with other contributors to risk and resilience.

## Data Availability

All data produced in the present study are available upon reasonable request to the authors

## ONLINE METHODS

### Whole-exome sequencing processing

#### Autism Spectrum Disorder (ASD)

We obtained the whole-exome sequencing (WES) data for 2,058 families from SSC^8, 35, 42^, including 1,597 quartets and 461 trios. These samples were generated from three centers (CSHL, UW, and YALE), and were sequenced on the Illumina HiSeq 2000 sequencing platform after being captured with the NimbleGen SeqCap EZ Exome v2 array. More information about this cohort can be found in Fischbach & Lord (2010)^89^.

#### Tourette Syndrome (TS)

WES data for 546 TS trios with male probands were derived from our previous work^34, 35^. In addition, we performed WES for another 24 newly recruited trios with male probands using the xGen Exome Research Panel (IDT) capture array and sequencing on the Hiseq 4000 platform. Any families with one or more reported Tic/TS affected parents were excluded from the analysis. Additional information about this cohort can be found in Wang *et al*., (2018)^35^.

#### Attention-deficit/hyperactivity disorder (ADHD)

We conducted whole-exome capture and sequencing for 341 ADHD male probands from the UK. Children and adolescents (aged 5 to 18 years) were recruited through Child and Adolescent Psychiatry or Pediatric outpatient UK clinics. ADHD diagnosis (according to DSM-III-R or DSM-IV) was confirmed using the parent version of the Child and Adolescent Psychiatric Assessment (CAPA)^90^, a semi-structured diagnostic interview. Samples with ASD were excluded. Approval for the study was obtained from the North West England and Wales Multicentre Research Ethics Committees. Written informed consent to participate was obtained from all parents and from adolescents aged 16-18 years old and assent was gained from children under 16 years of age. These samples were derived from primary blood cells and captured by the xGen Exome Research Panel (IDT) capture array and sequenced on the Hiseq 4000 platform. We did not generate parental sequencing data for these samples.

#### Epileptic Encephalopathy (EE)

We obtained WES data for 223 EE male probands from the Epi4K consortium. More information about these samples can be found in the published study^82^.

### Quality Control

We excluded samples with unexpected relationships by using a custom script based on PLINK^91^. Additionally, we excluded any samples with an inconsistent sex inferred from sex chromosome SNPs. After quality control, we obtained 1,975 ASD probands from the SSC (male:female = 1,661:314), 1,680 SSC siblings (male:female = 746:934), 570 male TS probands, 332 ADHD male probands, and 223 EE male probands. To avoid bias introduced by using proband-sibling pairs from the same family in burden analyses, we further removed 1) the male ASD proband in a quartet family with two male children, as the number of male unaffected siblings is smaller than male probands and therefore more limiting; and 2) the female unaffected sibling in a quartet family with two female children, as the sample size of female probands is much smaller than female controls. This step further excluded 647 male ASD probands (1,014 male ASD SSC probands remained) and 123 female SSC siblings (811 female SSC siblings remained).

In addition, we included an additional 22,416 male ASD probands and 1,638 male unaffected siblings from the SPARK pilot study as well as releases 1, 2, and 3. We used Hail 2.0 (https://github.com/hail-is/hail) to liftover the variants to GRCh37. We excluded samples with inconsistent sex information inferred from the impute_sex() function. The relationship between samples was inferred with identity_by_descent() and any unexpectedly related samples were removed. We further excluded any families without maternal samples available. As a result, we included 12,441 male probands and 1,621 male unaffected siblings with maternal samples in our study. Based on our model, i.e. the unaffected mother is a carrier of damaging variants penetrant mainly in males, we defined “model compatible” families as SPARK non-multiplex families (8,672 male probands and 1,355 male controls) as well as multiplex families that only have multiple affected male children (i.e. both parents are unaffected and no affected female children, 2,719 male probands and 194 male sibling controls). We added these samples based on the hypothesis that male probands in these families would be enriched for rare maternally-transmitted damaging variants within Chr X RERs. Indeed, we observed significant over-transmission of rare damaging variants (AF ≤ 1%) in RERs only in the SPARK model compatible sample set (OR 1.32, p = 0.010) and the rate of over-transmission is not significantly different from the rate in the SPARK non-multiplex sample set alone (OR 1.07, p = 0.33; **Figure S4**). In total therefore, we included 11,391 male ASD probands and 1,549 male unaffected siblings from the SPARK dataset.

### Family-based analysis

We utilized GATK Best Practices for data preprocessing and variant discovery^60^. To reduce potential batch effects in variant calling, we jointly genotyped all maternal samples across the SSC ASD datasets and applied a variant quality score (VQSR) filter to extract the variants passed VQSR. We further applied these filters to the called variants in maternal samples: 1) DP ≥ 20; 2) AB ≥ 0.3 and AB ≤ 0.7; 3) GQ ≥ 20. Instead of applying the VQSR, we conducted variant filtering separately for male probands/siblings and female probands/siblings using hard filters. More specifically, we required: 1) DP ≥ 10 for male children and DP ≥ 20 for female children; 2) AB ≥ 0.95 for male children, AB ≥ 0.3 and AB ≤ 0.7 for female children; 3) GQ ≥ 90. Again, we did not implement joint genotyping for female probands/siblings to make the analysis comparable to that in male probands/siblings. Likewise, we excluded paternally transmitted variants in female samples (i.e. the called variant should be heterozygous in mother and the offspring [like for male samples], but reference hemizygous in paternal sample). We excluded samples with an outlier number of rare variant calls (defined as mean ± 3 standard deviations).

For the SPARK samples, we could not use the same filters as above because 1) part of the VCF files from SPARK are post-filtered against VQSR; 2) variants in Chr X non-PAR in males are called diploid as well, which will affect the GQ estimation. Therefore, we adjusted our criteria as follows: 1) all variants should pass VQSR; 2) DP ≥ 20; 3) AB ≥ 0.95 for male children and AB ≥ 0.3 and AB ≤ 0.7 maternal samples [we did not analyze female children]; 4) GQ ≥ 20; 5) AF ≤ 0.1% in the maternal samples. Although these modifications potentially affect the sensitivity of the variant calling, the specificity remains the same as that in SSC data.

Within males, we labeled the variants that are maternal heterozygous and alternative hemizygous in the child as “transmitted” variants and variants that are maternal heterozygous variants but reference hemizygous in the child as “untransmitted”.

### Case-control analysis

To investigate whether the risk we observed for ASD in RERs exists in other male biased neurodevelopmental disorders, we conducted a similar analysis using TS and ADHD male probands. We also assessed EE male probands as a negative control, given that EE does not have a strong sex bias. As maternal samples were not available for all of the probands, we conducted case-control analyses.

We collected the VCF files for each individual and further applied the following filters on the callset: 1) DP ≥ 10; 2) AB ≥ 0.95; 3) GQ ≥ 90; 4) AF≤ 0.1% in our dataset as well as in ExAC v0.3 to generate the final call set. After calling, we excluded samples with an outlier number of rare variant calls (defined as mean ± 3 standard deviations). After applying these filters, we ended up with variant calls for 995 ASD SSC male probands and 730 SSC male siblings, 561 TS male probands, 329 ADHD male probands, and 220 EE male probands.

We estimated the variant calling accuracy for rare variants by comparing it with the SSC ASD trio-based (maternally-transmitted) variant calls and determined 97.24% recall and 91.13% precision. We applied a similar method with the full TS trios corresponding to the TS cases utilized here and obtained 96.90% recall and 91.15% precision. For ADHD samples, we attempted to confirm all the coding variants we identified on Chr X. In total, we assessed 260 variants by Sanger sequencing. Of the 236 variants with high quality sequencing data, 232 verified (98.3% confirmation rate). The additional 24 variants were putatively confirmed, although the sequencing results were noisy. Taken together, the confirmation rate in ADHD samples is nearly 90%, even if we conservatively posited that the 24 noisy sequence validations were “false positive” calls (232 / (236+24) = 89.23%), which is very close to our estimation of precision using the trio dataset.

### Population stratification

To ensure the results we observed were not driven by population stratification, we inferred the population ancestry for each dataset. We merged our dataset with genotypes from the 1000 Genomes Project and conducted the PCA analysis with the hail.pca() function. We then trained a random forest model using sklearn::RandomForestClassifier() on 1000 Genomes Project samples and predicted the ancestry of our samples. The majority of our samples were predicted to have European ancestry. When using only samples with European ancestry, rates of rare synonymous mutations are well-matched across samples from different datasets (**Figure S1B**).

### Identification of risk-enriched regions (RERs)

To identify RERs, we utilized microarray data from 65 families from the Simons Simplex Collection (SSC), each with one male proband and two or more male siblings. All the samples were genotyped on Illumina 1Mv1, 1Mv3, or Omni2.5 arrays and mapped to GRCh37 coordinates and have been reported in prior work^8^. We confirmed consistency between the reported sex and sex chromosome karyotype.

We first inferred the origins of Chr X (i.e. which maternal chromosome was inherited) for each of the children by assessing genotypes on either side of the centromere and selected the subset of families where at least one of the unaffected siblings shares the same Chr X origin as the proband (n = 48). Next, we identified SNPs of Chr X non-PAR that consistently segregated to the male probands. We then computed the density of ‘informative recombination’ events at each SNP across the 48 families, visualized the resulting chromosome-wide density plot, and identified four clear peaks. We extended 4.5 MB upstream and downstream from each peak to obtain four RERs that consistently segregate with risk (**Figure 1A**, GRCh37 coordinates: X:8625243-17625243, X:25181023-34181023, X:114438272-123438272, and X:136752585-145752585; see **Table S1** for a list of the 149 genes).

We compared the density curve with the published HapMap dataset^75^. The entire Chr X non-PAR was split into roughly 150 bins with a 1MB binwidth. We then calculated the average recombination rate (cM/MB) in each bin. We further normalized the average recombination rates by the sum of recombination rates of Chr X non-PAR in order to make a density distribution to compare with our dataset. For our dataset, we generated a comparable density distribution by counting the number of detected SNPs in each bin and normalizing the count by the total number of detected SNPs. We then compared the normalized values in our dataset with the normalized values from HapMap using a Wilcoxon signed rank test.

### Burden analysis

#### Burden analysis for rare transmitted LGD variants on Chr X non-PAR

To control for differences in ancestries, whole-exome capture platforms, and sequencing methods, we normalized LGD variants by synonymous variant counts with fisher.test(matrix(c(#LGD_case, #Syn_case, #LGD_ctrl, #Syn_ctrl), ncol = 2), alternative = “greater”). We further conducted the analysis using the same cutoff for allele frequency with AF ≤ 0.25% for the rare variants to make it comparable with the previous study by Lim *et al*^54^.

#### Burden analysis for rare transmitted damaging variants

We utilized rare synonymous variants to normalize the mutation rate in RERs and other NERs (non-enriched regions). Specifically, we performed Fisher’s exact test as fisher.test(matrix(c(#Dam_case, #Syn_case, #Dam_ctrl, #Syn_ctrl), ncol = 2), alternative = “greater”) to compare the rare transmitted mutation rate in male probands vs controls. We performed two additional analyses as negative controls: first, we conducted the same comparisons using more common variants (ExAC MAF>0.1%). Second, we performed the same analysis using female probands and unaffected sibling controls. We utilized rare variants that were inherited from the mother only (i.e. genotypes of a given site are heterozygous for daughter and mother and reference hemizygous in the father). Finally, we compared the mutation rates per individual directly in probands versus controls using a one-sided t-test and observed that the rare damaging variant rate is still elevated in male probands, thereby suggesting that enrichment of rare damaging variants within RERs is not driven by normalization via synonymous variants.

#### Burden analysis in the SPARK only, and SSC-SPARK combined dataset

To confirm the observation of enrichment of rare transmitted damaging variants in ASD probands, we utilized 11,391 male probands and 1,549 male controls from SPARK dataset to compare the transmission frequencies of rare variants in RERs and NERs. We calculated the odds ratio with a one-sided Fisher’s exact test:

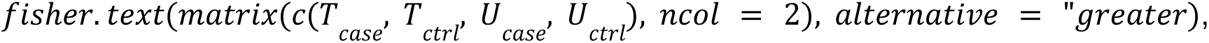

where T and U are transmitted and untransmitted variant counts, respectively. We performed this analysis for rare LGD, Mis3, and damaging (LGD + Mis3) variants and synonymous variants separately. Since this analysis is a family-based comparison, differences in population stratification and sequencing platforms are unlikely to confound the results.

We subsequently repeat this analysis after combining the SSC and SPARK dataset. Since the untransmitted variants in each individual serve as controls, we added back SSC probands to increase the statistical power in the transmission-disequilibrium test (cases: 1,661 SSC probands + 11,391 SPARK probands; controls: 746 SSC siblings + 1,549 SPARK controls).

Moreover, to investigate whether the over-transmission of rare damaging variants in ASD probands varies in terms of allele frequencies as well as family structures. We split the samples into three categories: (1) all families; 2) model compatible families, for which we (a) removed the families that are marked as “multiplex-multigenerational” or “multiplex-siblings-multigenerational” family type, (b) removed any families with ≥ 1 affected female sample; and (3) Non-multiplex families (i.e. removing any families that are labeled as “multiplex” for the family type). We aggregated the transmitted variant counts for damaging variants and synonymous variants under different allele frequency cutoffs (0.1%, 0.2%, 0.5%, 1%, 2%, 3%, 4%, 5%, 10%, 15%) and performed the transmission disequilibrium test with the above formula.

#### Burden analysis regarding for elevated ADHD symptoms

We categorized our samples into two groups based on whether the sample showed elevated ADHD symptoms. We defined any probands with an ADHD t-score of CBCL age 2-5 or 6-18 subscales greater than 70 as “ASD + ADHD” (i.e. ASD comorbid with ADHD), and the remainder as “ASD - ADHD” (i.e. ASD not comorbid with ADHD). We then performed the burden analysis in the two groups (150 probands in the “ASD + ADHD” group and 830 in the “ASD - ADHD” group). Direct comparisons between ASD with or without ADHD were conducted by comparing the rare damaging mutation rate after normalizing by synonymous variants with Fisher’s exact test.

We next performed a similar analysis to compare probands with different reported non-verbal IQs (nvIQs). We split the probands into two groups: lower nvIQ (nvIQ < 85, N = 593) and average or higher nvIQ (nvIQ ≥ 85, N = 389). Again, SSC unaffected siblings were used as controls. A direct comparison between nvIQ < 85 and nvIQ ≥ 85 was carried out by comparing the rare damaging mutation rate after normalizing by synonymous variants as described above.

#### Regression analysis of rare damaging variants in different cohorts

To understand the contribution of risk to different phenotypes, we performed a regression analysis using comorbidity status as a covariate. For the TS and ADHD cohorts, we used only the samples with clear records of clinical comorbidities. For the SSC cohort, we manually annotated samples as having elevated ADHD symptoms by setting a t-score cutoff of 70 from the CBCL age 2-5 or 6-18 subscales. Any probands with scores greater than the cutoff were considered to have elevated ADHD symptoms. We did not remove the small number of SSC siblings who had ADHD CBCL t-scores above the cutoff, as this results in a more conservative analysis. To improve statistical power, we excluded any groups with fewer than 100 samples. As a result, we obtained five groups of samples, including ASD only, TS only, ADHD only, ADHD + ASD, and ADHD + TS. TS comorbidity in SSC cohort is not well reported as we cannot distinguish whether the records are Tic disorder or TS. We conducted the Poisson regression analysis with the formula #dam ∼ Phenotype + offset(#syn), where #dam indicates the number of rare damaging variants in RERs. We used #syn, the rare synonymous count, as an offset to control for population ancestry and batch effects introduced by sequencing platforms. We further obtained the rate ratios, 95% confidence intervals, and p values.

### Comparison of RER genes and genes with high recombination rates

We inferred the recombination rates for all genes on Chr X non-PAR using recombination rates from the HapMap dataset^75^. We then selected the same number of genes with the highest recombination rate as the “top recombination rate genes”. We compared the “top recombination rate genes” with genes from RERs. Permutation testing was performed to estimate the overlap of the two sets of genes. We randomly selected the same number of genes from Chr X non-PAR and checked the overlap with the RER genes. We defined a “success” as the overlapped gene number being not less than the true overlap. We ran 100,000 permutations and obtained the p values #success / 100,000. The enrichment fold was calculated by the true overlapped gene number / mean(permuted overlapped gene number). We then utilized the genes that a) only existed in the top recombination rate gene set, b) only existed in RERs, and c) overlapped in the two sets of genes, to perform a damaging rare variant burden analysis (as described above).

### Estimation of the contribution of variants and identification of risk genes

We leveraged detected variants from the case-control dataset to estimate the contribution of damaging variants in RERs with estimated methods^34^. By utilizing SSC unaffected siblings as controls, we were able to infer the percent of probands with different disorder(s) who have at least one damaging variant in RERs, which we defined as the explainable rate by damaging RER variants. The 95% confidence interval was generated by bootstrapping. Furthermore, we were also able to estimate the percentage of damaging variants in RERs carrying risk by comparing the average mutation rate between cases and controls using t.test in R. The 95% confidence interval was obtained from this process directly. We applied similar methods to the variants that were detected from trios, and the results are very close to those presented here.

### Risk gene identification with SSC and SPARK samples

We utilized rare maternal-transmitted variants with allele frequency less than 0.1% from model compatible families to identify risk genes in the refined RERs (cases: 1,661 SSC probands + 11,391 SPARK probands; controls: 746 SSC siblings + 1,549 SPARK controls). Since we are lacking sufficient control samples/variants to estimate per gene transmission probabilities, we estimated the local transmission probability instead. Specifically, we split Chr X non-PAR into windows with 3MB size (so that each RER includes 3 windows) and estimated the transmission probability for each window with rare damaging variants from controls, and subsequently used it as the null transmission probability for genes included in the window. For genes that are included in two adjacent bins, we used the higher transmission probability for the risk gene identification (i.e. the more conservative estimate). We further excluded windows that are with less than 10 heterozygous variants in the mothers. The gene discovery analysis was performed with a one-sided binomial test for each gene:

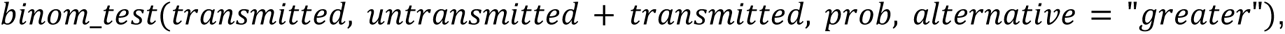

We then corrected the p values for multiple testing with the Bonferroni method at the risk regions level (n = 149, significant p = 0.05/149 = 0.00034), the Chr X non-PAR level (n = 808, significant p = 0.05/808 = 6.19E-5), and the exome-wide level (n = 19,251, significant p = 0.05/19,251 = 2.60E-6).

### Burden analysis of rare LGD variants from previous work

We applied the definition of RERs/NERs to samples from Lim et al. 2013^54^ to do a comparative burden analysis. In their dataset, we observed 12 LGD variants in RERs from male cases and 4 LGD variants in RERs from male controls (one-sided poisson test Rate Ratio 2.15, p = 0.13). In comparison, we found 48 and 24 LGD variants in NERs from male cases and controls, respectively (RR 1.43, p = 0.091). A comparison between RERs and NERs revealed a tendency of enrichment of rare LGD variants within RERs (OR 1.49, p = 0.37). Missense variants were not reported in this study.

Recently a study with larger samples published by Zhou et al.^46^ enabled us to directly evaluate whether the rare LGD variants are more likely to be transmitted in the male probands. We tabulated 155 transmitted LGD variants and 119 untransmitted variants in RERs and 698 transmitted and 671 untransmitted LGD variants in NERs. Burden analysis comparing RERs and NERs showed significant over-transmission of rare LGDs in RERs (OR 1.25, p = 0.05). Again, missense variants were not reported in this study.

## ACKNOWLEDGEMENTS

We wish to thank all of the families who have participated in this study, as well as all of the individuals involved in recruitment and assessment. We greatly appreciate rapid and generous access to published data from the SSC and SPARK datasets via SFARI Base (https://base.sfari.org); the Epi4K Consortium (https://www.epgp.org/) via Daniel Lowenstein, David Goldstein, and Erin Heinzen; the Tourette International Collaborative Genetics Study (TIC Genetics, https://tic-genetics.org/); the Tourette Syndrome Genetics Southern and Eastern Europe Initiative (TSGENESEE, http://tsgenesee.mbg.duth.gr/); the Tourette Association of America International Consortium for Genetics (TAAICG, https://tourette.org/); and the Uppsala Tourette Cohort (UTC). We also thank Sarah Pyle for graphic design, Vanessa Hus Bal for expert advice, Helen R. Willsey for unwavering support and scientific feedback, and the Willsey Lab along with our extended network of colleagues for their hard work and dedication.

This study was supported by grants from the National Institute of Mental Health to A.J.W and M.W.S (R01MH115963), A.J.W (R01NS105746), G.A.H. and J.A.T. (R01MH115958), Alyssa Rosen (R01MH115960), Donald L. Gilbert (R01MH115962), Samuel Kuperman (R01MH115961), Samuel H. Zinner (R01MH115993), and Barbara J. Coffey (R01MH115959); from the Tourette Association of America to A.J.W. (Young Investigator Award); from the Human Genetics Institute of New Jersey to G.A.H. and J.A.T.; and the New Jersey Center for Tourette Syndrome and Associated Disorders (NJCTS) to G.A.H. and J.A.T. We are also grateful to the NJCTS for facilitating the inception and organization of the TIC Genetics study. This study was also supported by the Weill Institute for Neurosciences (Startup Funding to A.J.W.) and the Overlook International Foundation (to M.W.S. and A.J.W.). The ADHD study was funded by the Wellcome Trust (to A.T., M.C.O., M.J.O.) and also by the MRC (A.T., K.L.), Action Research (A.T., M.C.O., M.J.O.) with project support from Sharifah Agha, Nigel Williams, Peter Holmans, and the core lab team at the MRC Centre for Neuropsychiatric Genetics and Genomics, Cardiff University. We also thank the Tourette Association of America International Consortium for Genetics (TAAICG) and the Tourette Syndrome Genetics Southern and Eastern Europe Initiative (TSGENESEE) for their ongoing collaboration and support. The content is solely the responsibility of the authors and does not necessarily represent the official views of the NIH or other funders.

## AUTHOR CONTRIBUTIONS

Conceptualization, SW and AJW; Methodology, SW, BW, and AJW; Software, SW and AJW; Validation, SW, VD, SD, NS, HA, JA, CD, and AJW; Formal Analysis, SW and AJW; Investigation, SW, BW, and AJW; Resources, TICGG, GAH, JAT, MWS, and AJW; Data Curation, SW, GAH, JAT, and AJW; Writing (Original Draft), SW, BW, MWS, and AJW; Writing (Review & Editing), SW, BW, NS, JA, CD, TICGG, JX, GAH, JAT, TVF, MCO, AT, MWS, and AJW; Visualization, SW and AJW; Supervision, AJW; Project Administration, GAH, JAT, AT, MWS, and AJW; Funding Acquisition, GAH, JAT, MWS, and AJW.

## COMPETING INTERESTS

Donald L. Gilbert has received salary/travel/honoraria from the Tourette Association of America, the Child Neurology Society, U.S. National Vaccine Injury Compensation Program, Ecopipam Pharmaceuticals, EryDel Pharmaceuticals, Elsevier, and Wolters Kluwer.

## SUPPLEMENTARY MATERIALS

### Figures

**Figure S1.**
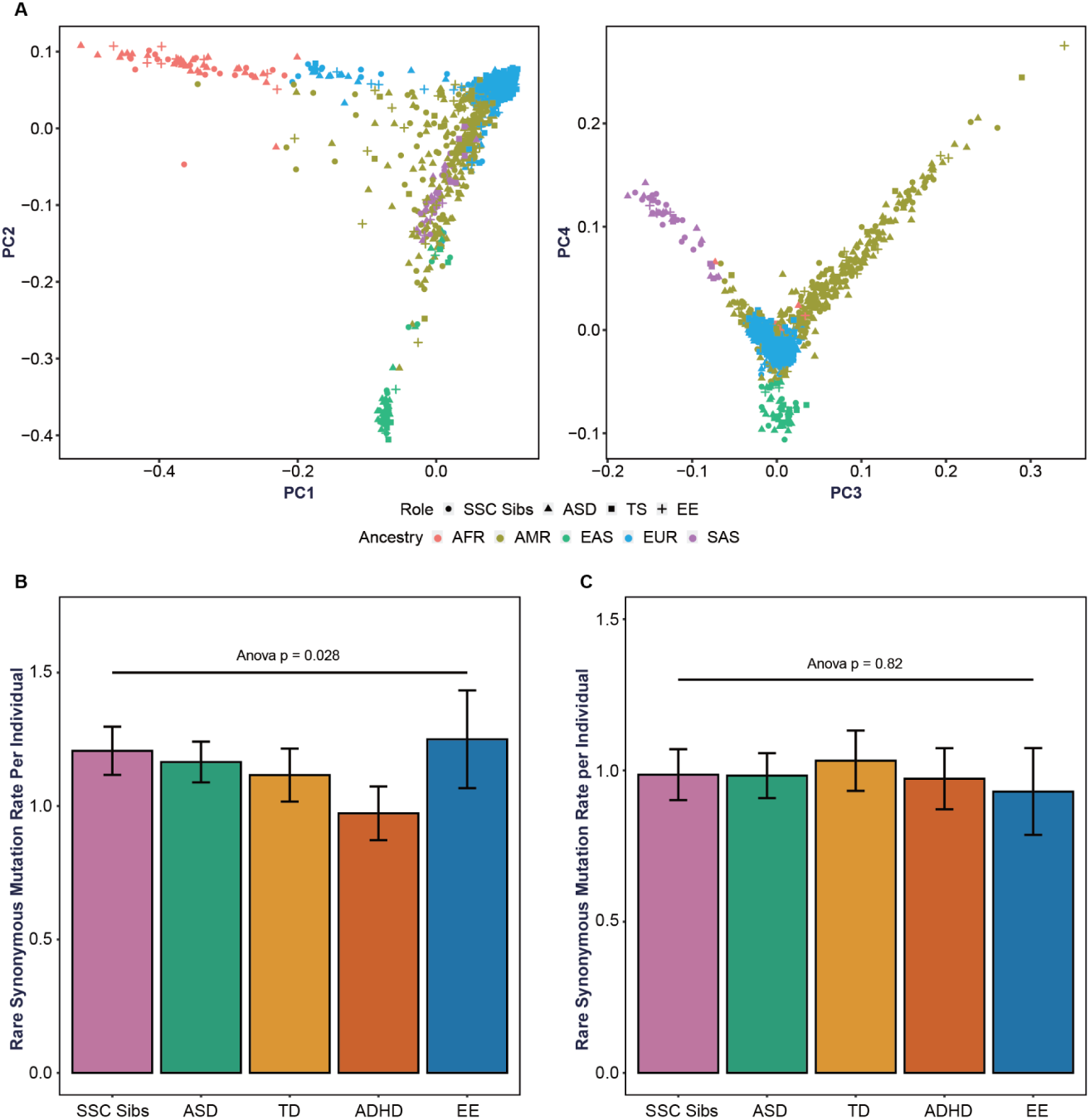
Population stratification by dataset and its impact on the rate of rare synonymous variants on Chr X non-PAR. (A) We performed PCA analysis for all the male samples in this study, and inferred the population composition with a random forest model, which was trained with data from the 1000 Genomes Project. The majority of our samples were inferred to have European ancestry. (B) To ensure that our variant calling pipeline does not lead to batch effects in different datasets, we compared the rare synonymous mutation rate per individual on Chr X non-PAR. We found that the mutation rates of rare variants from different datasets are significantly different across different datasets, which vary in population stratification. (C) However, when only using individuals from European ancestry, the rare synonymous mutation rates are similar across each dataset, suggesting that (1) our variant pipeline does not appear to be introducing biases across datasets and that controlling by synonymous rate should control for differences in ancestry (see also **Figure S3**).

**Figure S2.**
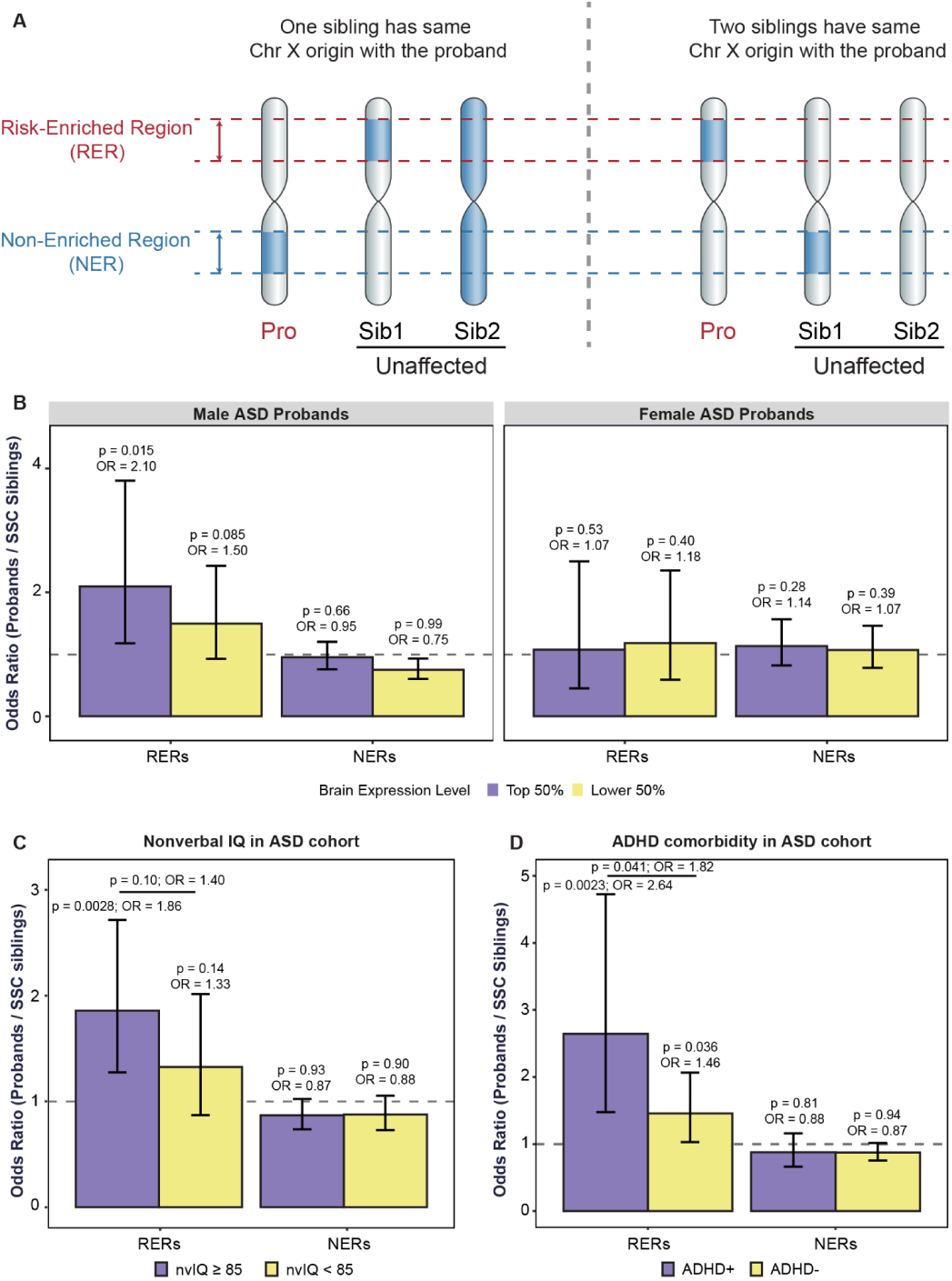
The rate of rare damaging variants per individual is elevated in RERs in probands only. (A) Schema showing how we derived risk-enriched regions (RERs) from microarray genotyping data.We included the families with at least three male children and only retained families with at least one unaffected sibling sharing the same Chr X origin as the proband (N = 65). We then identified regions within the Chr X non-pseudoautosomal regions (non-Par) that consistently segregated with ASD status and termed these RERs. We then termed any regions outside RERs, but within Chr X non-PAR, as non-risk-enriched regions (RERS). (B) Rare transmitted damaging variants tend to be more enriched in RER genes with higher brain expression in males. Gene expression levels were estimated from BrainSpan^70, 71^.(C) The probands were grouped into two based on the median of non-verbal IQ (nvIQ) in the ASD probands. Although the difference between the two groups is not significant in RERs, the probands with higher nvIQ (nvIQ ≥ 85) tend to have a higher signal. (D) We categorized ASD probands into probands with comorbid ADHD (ADHD+) and without comorbid ADHD (ADHD-). While both groups show significant enrichment of rare damaging variants in RERs, the mutation rate in the ADHD+ group is significantly higher.

**Figure S3.**
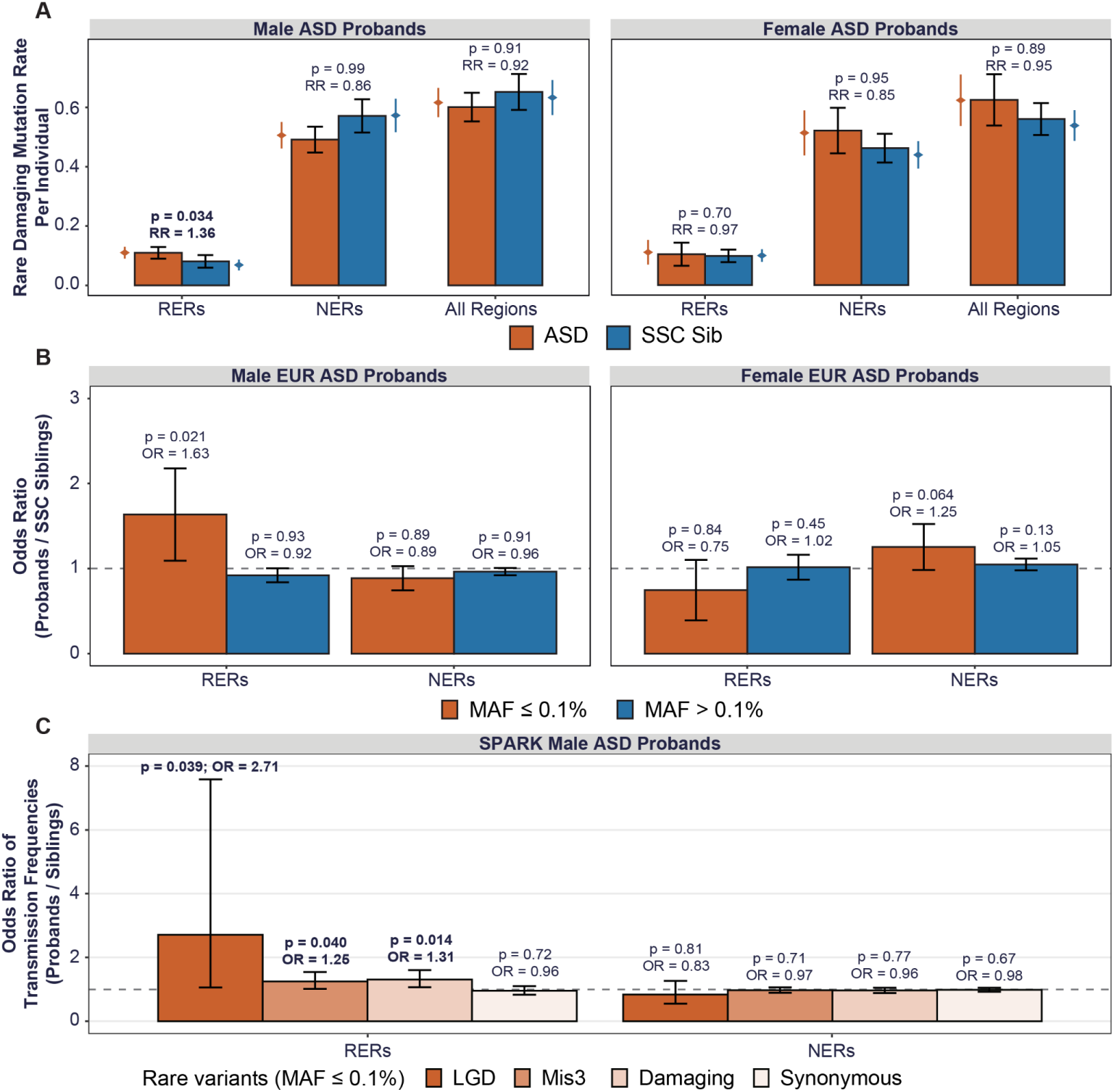
Rare transmitted damaging mutation rate is significantly increased in male ASD probands. (A) We compared the rate of rare damaging mutations per individual between ASD probands and SSC siblings, for males and females separately (one-sided t-tests; bar plots; error bars denote 95% confidence intervals). We also show the rate normalized by rare synonymous mutations (red and blue “points” on either side of the bar plots; these are the values compared in the main text). Again, error bars denote 95% confidence intervals. In both cases, the rate of rare damaging mutations is specifically increased in RERs in male probands only, suggesting that normalization by the rate of synonymous variants is not driving our observation of a significant enrichment of rare damaging variants within RERs. (B) Given the population stratification and differences in synonymous variant rates observed in Figure S1, we conducted the main text burden analysis using European samples only (comparing the rate of rare damaging variants normalized by the rate of synonymous variants). Again, we observed significant enrichment for rare damaging variants in RERs from male probands only. (C) We conducted a burden analysis to investigate whether transmission frequencies of different mutations are elevated in male ASD probands from SPARK dataset. Compared to male controls, rare LGD, Mis3, and damaging (LGD+Mis3) variants are all significantly over-transmitted to the male ASD probands.

**Figure S4.**
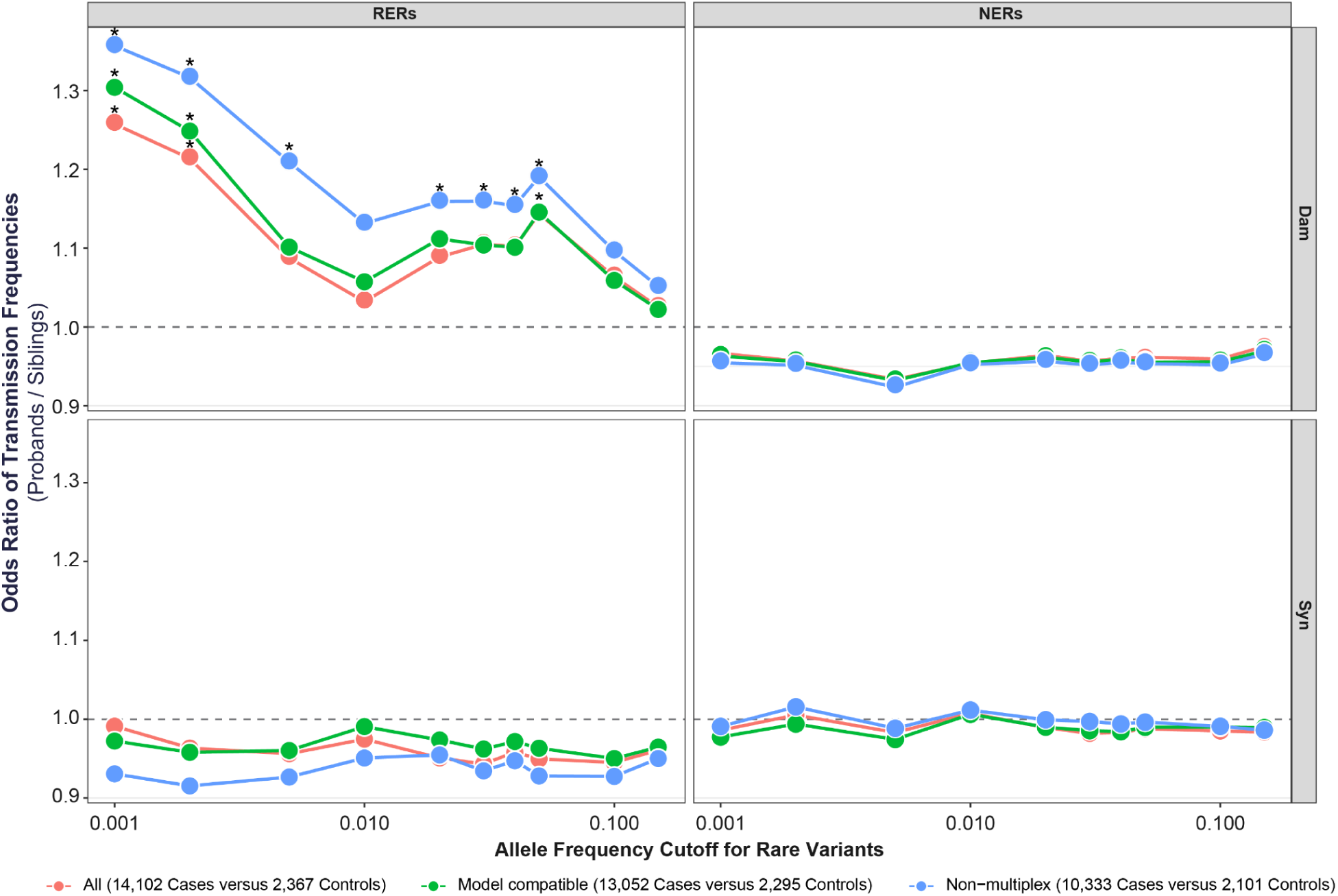
Rare damaging variants are over-transmitted to probands in risk-enriched regions only. We filtered rare variants based on different allele frequency thresholds and then conducted burden analyses with SSC and SPARK samples. We repeated this analysis with three different datasets: all male samples, regardless of the family type (All); male samples from SSC (simplex) families, non-multiplex SPARK families, and non-multi-generational multiplex SPARK families with only male affected children (Model compatible); and all male samples but excluding all multiplex families (Non-multiplex). Asterisks indicate significant burden analyses(uncorrected p < 0.05).

### Tables

**All tables are included in a separate file (Table S1-4).**

**Table S1. See separate excel file.**

Table S1-1. Trio samples included in the analysis

Table S1-2. Case-control samples included in the analysis (Male only)

Table S1-3. Risk gene identification with SSC and SPARK datasets

**Table S2.** Rare transmitted LGD variants are enriched in ASD male probands. We conducted a burden analysis for all the rare transmitted variants in Chr X non-PAR or inside the risk-enriched regions (RERs) with a one-sided fisher’s exact test. LGD: likely gene-disrupting; Mis3: missense variants with PolyPhen2 [HDIV] score ≥ 0.957; Damaging: LGD + Mis3. To reproduce the previous study^54^, in addition to the threshold of the rare variants used throughout our study (AF ≤ 0.1%), we included AF ≤ 0.25% in this table to more accurately compare our results to Lim *et al*^54^. ^1^ AF ≤ 0.25% in our dataset as well as ExAC v0.3; ^2^ AF ≤ 0.1% in our dataset as well as ExAC v0.3

|  | Overall in Chr X non-PAR |  |  |  |  |  |  |  | RERs |  |
| --- | --- | --- | --- | --- | --- | --- | --- | --- | --- | --- |
|  | LGD (AF≤0.25%<sup>1</sup>)<br>(Pro versus Sib) |  | LGD (AF≤0.1%<sup>2</sup>)<br>(Pro versus Sib) |  | Mis3 (AF≤0.1%)<br>(Pro versus Sib) |  | Damaging (AF≤0.1%)<br>(Pro versus Sib) |  | Damaging (AF≤0.1%)<br>(Pro versus Sib) |  |
|  | OR<br>(95% CI) | p-val | OR<br>(95% CI) | p-val | OR<br>(95% CI) | p-val | OR<br>(95% CI) | p-val | OR<br>(95% CI) | p-val |
| **Male** | 1.78<br>(1.11 - 2.91) | 0.019 | 1.86<br>(1.08 - 3.30) | 0.028 | 0.94<br>(0.83 - 1.08) | 0.78 | 0.97<br>(0.86 - 1.11) | 0.65 | 1.60<br>(1.15 - 2.24) | 0.0084 |
| **Female** | 1.19<br>(0.65 - 2.11) | 0.35 | 1.13<br>(0.50 - 2.42) | 0.46 | 1.16<br>(0.96 - 1.39) | 0.095 | 1.16<br>(0.97 - 1.39) | 0.093 | 1.11<br>(0.69 - 1.77) | 0.39 |

**Table S3.** Rare LGD variants in male probands. We conducted a burden analysis for all the rare variants in Chr X non-PAR or inside the risk-enriched regions (RERs) with a one-sided fisher’s exact test. LGD: likely gene-disrupting; Mis3: missense variants with PolyPhen2 [HDIV] score ≥ 0.957; Damaging: LGD + Mis3. To reproduce the previous study^54^, in addition to the threshold of the rare variants throughout our study (AF ≤ 0.1%), we include AF ≤ 0.25% in this table to more accurately compare our results to Lim *et al*^54^.

|  | Overall in Chr X non-PAR |  |  |  |  |  |  |  | RERs |  |
| --- | --- | --- | --- | --- | --- | --- | --- | --- | --- | --- |
|  | LGD (AF≤0.25%)<br>(Pro versus Sib) |  | LGD<br>(Pro versus Sib) |  | Mis3<br>(Pro versus Sib) |  | Damaging<br>(Pro versus Sib) |  | Damaging<br>(Pro versus Sib) |  |
|  | OR<br>(95% CI) | p-val | OR<br>(95% CI) | p-val | OR<br>(95% CI) | p-val | OR<br>(95% CI) | p-val | OR<br>(95% CI) | p-val |
| **ASD** | 1.34<br>(0.90 - 2.03) | 0.12 | 1.35<br>(0.85 - 2.18) | 0.15 | 0.92<br>(0.81 - 1.04) | 0.88 | 0.94<br>(0.83 - 1.06) | 0.82 | 1.45<br>(1.05 - 2.00) | 0.027 |
| **TS** | 1.58<br>(1.00 - 2.49) | 0.047 | 1.53<br>(0.90 - 2.59) | 0.096 | 1.01<br>(0.87 - 1.17) | 0.48 | 1.03<br>(0.89 - 1.19) | 0.38 | 2.12<br>(1.46 - 3.08) | 0.00032 |
| **ADHD** | 1.77<br>(1.04 - 2.98) | 0.041 | 1.91<br>(1.04 - 3.46) | 0.039 | 1.14<br>(0.96 - 1.37) | 0.11 | 1.18<br>(0.99 - 1.41) | 0.062 | 2.55<br>(1.60 - 4.08) | 0.00032 |
| **EE** | 0.60<br>(0.24 - 1.31) | 0.89 | 0.56<br>(0.18 - 1.44) | 0.91 | 1.01<br>(0.83 - 1.23) | 0.48 | 0.99<br>(0.82 - 1.20) | 0.55 | 1.11<br>(0.65 - 1.87) | 0.41 |

**Table S4.** Rare maternally-transmitted synonymous variants on Chr X are more undercalled than rare maternally-transmitted synonymous variants on the autosomes. We detected rare maternally-transmitted synonymous variants in SPARK male samples on the autosomes and on Chr X using the same criteria described in the Methods. For autosomal variants, the transmitted variants were defined as reference homozygous in father, heterozygous in mother, and heterozygous in child, while the untransmitted variants were defined as reference homozygous in father, heterozygous in mother, and reference homozygous in child. For Chr X variants, the transmitted variants were defined as heterozygous in mother, and alternative hemizygous in child, while the untransmitted variants were defined as heterozygous in mother, and reference hemizygous in child.

| Chromosome | Role | Number Transmitted | Number Untransmitted | Transmission frequency |
| --- | --- | --- | --- | --- |
| Chr X | Case | 11431 | 13297 | 0.462269 |
|  | Ctrl | 1743 | 1980 | 0.468171 |
| Autosomes<br>(Chr 1-22) | Case | 196547 | 204771 | 0.489754 |
|  | Ctrl | 44777 | 45649 | 0.495178 |

## Notes

### Author Declarations

Human Research Protection Program Institutional Review Board (IRB) of University of California, San Francisco gave ethical approval for this work.

