## Supplemental Note for "Rare Maternally Inherited Coding Variants on Chromosome X Carry Predominantly Male Risk in Autism, Tourette Syndrome, and Attention-deficit/Hyperactivity Disorder"

**Consortium Author List**
**Tourette International Collaborative Genetics (TIC Genetics)**

Mohamed Abdulkadir[1], Hasan Alkhairo[2], Juan Arbelaez[2], Vanessa H. Bal[3], Yana Bromberg[4], Lawrence W. Brown[5], Xiaolong Cao[6], Keun-Ah Cheon[7], Barbara J. Coffey[8], Li Deng[6], Andrea Dietrich[9], Sam Drake[2], Vanessa Drury[2], Clif Duhn[2], Thomas V. Fernandez[10], Carolin Fremer[11], Blanca Garcia-Delgar[12], Donald L. Gilbert[13], Danea Glover[6], Dorothy E. Grice[14], Tammy Hedderly[15], Gary A. Heiman[6], Isobel Heyman[16], Pieter J. Hoekstra[9], Hyun Ju Hong[17], Chaim Huyser[18], Eunjoo Kim[19], Young Key Kim[20], Young-Shin Kim[21], Robert A. King[22], Yun-Joo Koh[15], Sodahm Kook[23], Samuel Kuperman[24], Kate Langley[25], Bennett L. Leventhal[21], Kirsten Müller-Vahl[11], Alexander Münchau[26], Marcos Madruga-Garrido[27], Joanna Martin[28], Dararat Mingbunjerdsuk[29], Pablo Mir[30], Astrid Morer[31], Tara L. Murphy[16], Cara Nasello[6], Michael C. O'Donovan[28], Michael J. Owen[28], Kerstin J. Plessen[30], Veit Roessner[32], Eun-Young Shin[33], Dong-Ho Song[34], Jungeun Song[35], Matthew W. State[2], Nawei Sun[2], Joshua K. Thackray[6], Anita Thapar[28], Jay A. Tischfield[6], Frank Visscher[36], Belinda Wang[2], Sheng Wang[2], A. Jeremy Willsey[37], Jinchuan Xing[6], and Samuel H. Zinner[38]

[1]University of Groningen, University Medical Center Groningen, Department of Child and Adolescent Psychiatry, Groningen, The Netherlands. 2. Rutgers, the State University of New Jersey, Department of Genetics and the Human Genetics Institute of New Jersey, Piscataway, NJ, USA.

[2]Department of Psychiatry and Behavioral Sciences, UCSF Weill Institute for Neurosciences, University of California, San Francisco, San Francisco, CA 94143, USA.

[3]Graduate School of Applied and Professional Psychology, Rutgers University, New Brunswick, New Jersey, USA.

[4]Department of Biochemistry and Microbiology, Rutgers, New Brunswick, NJ.

[5]ChildrenÕs Hospital of Philadelphia, Philadelphia, PA, USA.

[6]Department of Genetics and the Human Genetics Institute of New Jersey, Rutgers, the State University of New Jersey, Piscataway, NJ, USA.

[7]Division of Child and Adolescent Psychiatry, Department of Psychiatry Institute of Behavioral Science in Medicine Yonsei University College of Medicine, Severance Hospital.

[8]Child and Adolescent Psychiatry, University of Miami Miller School of Medicine, Miami, FL, USA.

[9]1) University of Groningen, University Medical Center Groningen, Department of Child and Adolescent Psychiatry, Groningen, The Netherlands 2) Accare Child Study Center, Groningen, the Netherlands.

[10]Yale Child Study Center and Department of Psychiatry, Yale University School of Medicine, New Haven, CT, USA.

[11]Clinic of Psychiatry, Socialpsychiatry and Psychotherapy, Hannover Medical School, Hannover, Germany.

[12]Department of Child and Adolescent Psychiatry and Psychology, Institute of Neurosciences, Hospital Clinic Universitari, Barcelona, Spain.

[13]Cincinnati Children's Hospital Medical Center, Cincinnati, OH, USA.

[14]Icahn School of Medicine at Mount Sinai, New York, NY, USA.

[15]Evelina London Children's Hospital GSTT, Kings Health Partners AHSC, London, UK.

[16]Great Ormond Street Hospital for Children, and UCL Institute of Child Health, London, UK.

[17]Hallym University Sacred Heart Hospital, Anyang, South Korea.

[18]1) Amsterdam UMC, Department of Child and Adolescent Psychiatry, Amsterdam, The Netherlands; 2)Levvel, Academic Center for Child and Adolescent Psychiatry, Amsterdam, The Netherlands.

[19]Gangnam Severance Hospital, Yonsei University College of Medicine, Gangnam-Gu Seoul, South Korea.

[20]Yonsei Bom Clinic, Seoul, South Korea.

[21]University of California, Department of Psychiatry, San Francisco, USA;.

[22]Yale Child Study Center and Department of Psychiatry, Yale University School of Medicine, New Haven, CT, USA.

[23]Gangbuk Samsung Hospital Workplace Mental Health Institute, Seoul, Korea.

[24]University of Iowa Carver College of Medicine, Iowa City, IA USA.

[25]MRC Centre for Neuropsychiatric Genetics and Genomics, Division of Psychological Medicine and Clinical Neurosciences, Cardiff University School of Medicine, Cardiff, Wales, United Kingdom School of Psychology, Cardiff University School of Medicine, Cardiff, Wales, United Kingdom.

[26]Institute of Systems Motor Science, Center of Brain, Behavior and Metabolism, University of Lübeck, Lübeck, Germany.

[27]Sección de Neuropediatría, Instituto de Biomedicina de Sevilla (IBiS), Hospital Universitario Virgen del Rocío/CSIC/Universidad de Sevilla, Seville, Spain.

[28]MRC Centre for Neuropsychiatric Genetics and Genomics, Division of Psychological Medicine and Clinical Neurosciences, Cardiff University School of Medicine, Cardiff, Wales, United Kingdom.

[29]Department of Neurology, University of Washington School of Medicine, Seattle, WA, USA.

[30]1) Unidad de Trastornos del Movimiento. Instituto de Biomedicina de Sevilla (IBiS). Hospital Universitario Virgen del Rocío/CSIC/Universidad de Sevilla. Seville, Spain; 2) Centro de Investigación Biomédica en Red sobre Enfermedades Neurodegenerativas (CIBERNED), Madrid, Spain.

[31]1) Department of Child and Adolescent Psychiatry and Psychology, Institute of Neurosciences, Hospital Clinic Universitari, Barcelona, Spain; 2) Institut d'Investigacions Biomediques August Pi i Sunyer (IDIBAPS), Barcelona, Spain; 3) Centro de Investigacion en Red de Salud Mental (CIBERSAM), Instituto Carlos III, Spain 4) University of Barcelona.

[32]Department of Child and Adolescent Psychiatry, Faculty of Medicine of the TU Dresden, Dresden, Germany.

[33]Yonsei Yoo & Kim Mental health clinic, Seoul, South Korea.

[34]Yonsei University Severance Hospital, Seoul, South Korea.

[35]National Health Insurance Service Ilsan Hospital, Goyang-si, South Korea.

[36]Admiraal De Ruyter Ziekenhuis, Department of Neurology, Goes, The Netherlands.

[37]Department of Psychiatry and Behavioral Sciences, UCSF Weill Institute for Neurosciences, University of California, San Francisco, San Francisco, CA 94143, USA Quantitative Biosciences Institute (QBI), University of California, San Francisco, San Francisco, CA 94143, USA.

[38]Department of Pediatrics, University of Washington School of Medicine, Seattle, WA, USA.
